# From Spectral Resolution to Speech Perception: A Review of Findings in Postlingually Deafened Adult Cochlear Implant Listeners

**DOI:** 10.1101/2025.04.28.25326599

**Authors:** Samin Ashjaei, Reed Farrar, Laura Droege, Madison Paxton, Kathryn Morgan, Meisam Arjmandi

## Abstract

**Purpose:** Reduced spectral resolution limits speech recognition in cochlear implant (CI) listeners. Although many studies have examined this association, uncertainties remain regarding its strength and contributing methodological and clinical factors. This narrative review synthesizes findings from studies of postlingually deafened adult CI listeners, focusing on psychophysical measures of spectral resolution and their strengths and limitations.

**Method:** We reviewed studies published through January 2025 that examined the relationship between psychophysical measures of spectral resolution and speech perception outcomes in postlingually deafened adult CI listeners. Twenty-four studies met inclusion criteria and tested this association statistically. Where available, the coefficient of determination (*R²*) was extracted to quantify the variance in speech recognition outcomes explained by spectral resolution measures.

**Results:** Several studies found a statistically significant association between psychophysical measures of spectral resolution and speech recognition performance. The strength of this association varied widely (*R²* = 0.21 to 0.68), depending on the spectral resolution measure and the speech material used. Variability in *R²* values reflects differences in test procedures, study populations, and speech materials.

**Conclusions:** Several psychophysical measures of spectral resolution are promising predictors of speech recognition and may serve as valuable tools for evaluating new CI signal processing algorithms, programming strategies, and auditory rehabilitation. A deeper understanding of the spectral resolution–speech perception relationship requires examining the distinct contributions of both peripheral and central auditory processes. Variability in observed associations highlights the need for further mechanistic research into the pathways linking spectral resolution to speech recognition outcomes.

## 1. Introduction

Cochlear implants (CIs) are the most effective neural prostheses for individuals with moderate to profound sensorineural hearing loss who gain no-to-limited benefit from hearing aids (e.g., Svirsky et al., 2000; Holden et al., 2013; Caswell-Midwinter et al., 2022). CI listeners experience significant improvements in hearing, speech, and language outcomes (e.g., Svirsky et al., 2000; Holden et al., 2013; Caswell-Midwinter et al., 2022; Arjmandi et al., 2022a). However, challenges persist, including large individual variability in outcomes that remains only partially explained, and significant difficulties with speech understanding in noise (e.g., Svirsky et al., 2000; Holden et al., 2013; Caswell-Midwinter et al., 2022; Arjmandi et al., 2022a, 2025, Ashjaei et al., 2022, 2024). Spectral resolution ability is a key focus in CI research and one of the most critical abilities for effective speech perception, particularly in noisy environments (e.g., O’Neill et al., 2019; Fu & Nogaki, 2005; Won et al., 2007; Friesen et al., 2001). It refers to listeners’ abilities to resolve spectral components of complex acoustic signals such as speech (e.g., Won et al., 2007; Litvak et al., 2007). Speech signals contain numerous co-varying acoustic cues, including spectral features such as formant frequencies (primarily the first and second formants; e.g., Sagi & Svirsky, 2017; Arjmandi et al., 2018), formant transitions (which also have a temporal component; e.g., Hedrick & Carney, 1997), spectral tilt (e.g., Alexander & Kluender, 2008), spectral cues for place of articulation (e.g., Dorman & Loizou, 1996), and the spectral composition of the release burst (e.g., Repp, 1984) in phonemes. The ability to perceptually distinguish between speech phonemes that differ in frequency information is essential for speech perception (e.g., Winn et al., 2016; DiNino et al., 2016; Nie, et al., 2005; Arjmandi et al., 2022a).

Studies on speech perception in CI listeners suggest that limited access to spectral contrasts accounts for a significant proportion of word and phoneme recognition errors (e.g., DiNino et al., 2016; Cucis et al., 2021; Winn et al., 2016; Arjmandi et al., 2025). Improving access to spectral cues may support better speech recognition outcomes in CI listeners, but this requires a clear understanding of how variations in spectral resolution affect speech perception. This knowledge helps clarify the extent to which difficulty in speech perception is due to limited spectral resolution, how effectively different measures predict speech outcomes, and how likely outcomes can be estimated for individual patients to better guide rehabilitation. Different methods of measuring spectral resolution may serve distinct purposes such as evaluating the effectiveness of new implant or electrode designs or signal processing strategies, identifying poorly functioning electrodes to fine-tune CI implant programming (e.g., using current focusing, current steering, or modified frequency-to-electrode allocation strategies etc.), and assessing the benefits of auditory trainings. Understanding these relationships allows us to select the most appropriate measures based on specific clinical or research goals and to guide the development of targeted strategies for improving outcomes.

The primary objective of this narrative review is to synthesize existing findings on the relationship between spectral resolution ability and speech recognition outcomes in postlingually deafened adult CI listeners. We examine results obtained in both quiet and noisy listening conditions, highlight consistent and divergent findings, discuss methodological challenges in establishing this relationship, and offer practical recommendations for future research. To assess the strength of this relationship, we report the coefficient of determination (*R²*) for each study, which quantifies the proportion of variance in speech perception explained by a measure of spectral resolution. Additionally, we review various speech perception measures used across studies to evaluate their effectiveness in establishing this relationship.

### 1.1. Potential Factors Explaining the Link Between Spectral Resolution and Speech Perception in CI Listeners

This review is centered on synthesizing findings related to the link between spectral resolution and speech perception in postlingually deafened CI listeners. Although not the primary focus, we provide a concise overview of key constructs, factors, and processes that may shape this relationship, solely to contextualize our discussion. One factor is the number of active CI electrodes, which may impact both speech perception and spectral resolution ability. CIs replace thousands of hair cells on the typical human cochlea with only 12 to 22 electrodes (e.g., Zeng et al., 2008; Loizou, 1998; O’Donoghue, 2013; Dorman & Wilson, 2004; Svirsky, 2017; Goupell, 2015), leading to a coarse representation of the sound spectrum and limitations in distinguishing between closely spaced frequency components (i.e., harmonics) (e.g., Shannon, 1995; Goupell, 2015; Henry, & Turner, 2003; Friesen et al., 2001; Holmes et al., 1987; Cychosz et al., 2024). The electrode number is inherently limited by physical space within the scala media of the cochlea (e.g., Goycoolea et al., 1990). Combined with this limited number of electrodes, there is a likelihood of current spread or channel interaction, where electrical stimulation from one electrode overlaps with adjacent electrodes, blurring the distinct spectral cues in speech and making it harder to perceive fine frequency differences for speech perception (e.g., Goehring et al., 2021; Jones et al., 2013; Cucis et al., 2021; Bierer & Litvak, 2016; Abbas et al., 2004).

The representation of spectral information through CI and its impact on speech perception can also be influenced by *CI mapping parameters*. For example, monopolar stimulation strategy creates broader electrical fields compared to bipolar stimulation strategy, which could increase channel interaction (e.g., Berenstein et al., 2008; Zhu et al., 2012; Kwon et al., 2006), reduce spectral resolution (e.g., Jones et al., 2013) and speech recognition outcomes (e.g., Cucis et al., 2021; Throckmorton et al., 2002). *CI speech coding strategies* could also impact spectral resolution and speech perception depending on how they encode spectral information in speech (e.g., Wouters et al., 2015; Liepins et al., 2020). Envelope-based strategies, such as continuous interleaved sampling (CIS) and advanced combination encoder (ACE), use a spectral encoding approach based on the function of auditory filter bank and envelope detection, which are limited in representing spectral fine structure cues in speech. In contrast, strategies like fine structure processing (FSP) and HiRes120 aim to transmit fine spectral cues (e.g., Wouters et al., 2015). These differences can affect the accessibility of frequency information for CI listeners, particularly influencing speech perception in noise (e.g., Wouters et al., 2015). Other factors of CI mapping, including pulse width (e.g., Skinner et al., 2002; Henry & Turner, 2003), stimulation rate (e.g., Azadpour et al., 2018), and dynamic range (e.g., Loizou & Poroy, 2001), can also impact spectral resolution and consequently, speech perception, although findings have been inconsistent.

The quality by which individual CI electrodes stimulate their target auditory nerves in the cochlea is defined as the *electrode-neuron interface (ENI) quality* (e.g., Arenberg Bierer, 2010; Arjmandi et al., 2022a). It is identified as a key determinant of spectral resolution as it can cause excessive channel interaction (e.g., Arjmandi et al., 2022a; DiNino et al., 2019; Arenberg Bierer, 2010; DeVries & Arenberg, 2018a). *Electrode placement* with deeper insertion depth and closer proximity to the modiolus are suggested to improve spectral fidelity and speech perception (e.g., Wanna et al, 2014; DeVries & Arenberg, 2018b). A reduced *electrode-neuron distance* provides more focused stimulation to the auditory nerve and decreases the spread of the neural excitation, which in turn increases the frequency specificity and spectral resolution (e.g., DeVries & Arenberg, 2018b; Long et al., 2014; Goldwyn et al., 2010; Jahn & Arenberg, 2020). Another dimension of ENI quality is *neural health*, which involves the survival and health of the auditory nerve fibers (e.g., Goldwyn et al., 2010; Jahn & Arenberg, 2020). Even when electrodes are positioned near the target region in the cochlea, a low number of surviving or healthy neurons may prevent the effective transmission of spectral information. The electrical current could spread beyond the targeted neural population due to the tissue growth and ossification, which impacts the impedance of each channel. This current spread reduces the ability to stimulate discrete neural regions, which may contribute to poorer spectral resolution and reduced speech perception.

In addition, CI’s frequency allocation map may not perfectly match the natural tonotopic organization of the cochlea, which leads to *frequency-place mismatch*. This misalignment can distort spectral cues, particularly in cases of shallow electrode insertion or anatomical differences in cochlear length (e.g., Xu et al., 2020; Canfarotta et al., 2020). *Electrode array design* may help mitigate challenges related to channel interaction and residual hearing preservation by improving electrode placement relative to the auditory nerves. For example, *perimodiolar arrays* position electrodes near the modiolus, *lateral wall arrays* align them along the outer wall of the cochlear tympani, and *mid-scala arrays* situate them centrally within the scala tympani. Deeper insertion, although within the safe limits to avoid traumas, covers a wider range of frequencies along the basilar membrane, which can potentially reduce the risk of frequency-place mismatch (e.g., Zeng et al., 2008). Despite these arguments, the current evidence regarding the effects of these factors on spectral resolution and speech perception in CI listeners is not yet compelling.

The relationship between spectral resolution ability and speech perception outcomes can be further influenced by *individual differences in central auditory processing* of spectral information and speech perception (e.g., Abbas et al., 2004; Scheperle & Abbas 2015a; Scheperle & Abbas 2015b; see also Farrar et al., 2024 for a review). This involves how the central auditory system interprets incoming auditory information from CI electrodes, including its ability to process, differentiate, and integrate frequency information, which may either facilitate (compensatory role) or hinder speech understanding. Even when peripheral factors like the CI mapping and ENI quality are optimized to some extent, individual variability in how the central auditory system processes incoming electrical signals can lead to differing perceptual outcomes (e.g., Scheperle & Abbas 2015a; Scheperle & Abbas 2015b; Farrar et al., 2024). The relationship between spectral resolution ability and speech perception can also be influenced by audiological factors including etiology of hearing loss, age at implantation, pre-implantation duration of auditory deprivation, duration of CI use, residual hearing, and receiving auditory rehabilitation services (e.g., Green et al., 2007 for a review on these factors). These factors may directly or indirectly affect neural health (e.g., Otte et al., 1978; Leake & Hradek, 1988), tissue growth or ossification (e.g., Kamakura & Nadol, 2016; Wilk et al., 2016), and central auditory processing of spectral information and its connection to speech recognition outcomes. For instance, higher age at implantation can negatively impact spectral resolution (e.g., Landsberger et al., 2018; Gifford et al., 2018), which could be due to reduced neural plasticity and increased neural degeneration. Prolonged under-stimulation of the auditory nerve fibers (long duration of haring loss) can exacerbate the degeneration of auditory nerves (e.g., Otte et al., 1978; Leake & Hradek, 1988) and central auditory nervous system, which could limit the brains’ ability to distinguish spectral components important for speech perception.

### 1.2. Spectral Resolution and Speech Recognition in Simulated CI Processing

Studies also used speech perception scores from listeners with typical hearing (TH) in response to stimuli with manipulated spectral information to simulate how spectral information is transmitted through CI signal processing algorithms. Spectral resolution can be manipulated using noise vocoders (e.g., Shannon et al., 1995; Friesen et al., 2001; Xu et al., 2005), sine vocoders (e.g., Dorman & Loizou, 1997), gaussian envelope tone vocoders (e.g., Goupell et al., 2013), and other methods to simulate spectral smearing (e.g., ter Keurs et al., 1993). After generating vocoded signals, changing the number of spectral channels, level of channel interaction, and electrode array insertion depth can simulate the spectral resolution ability in CI listeners (see Cychosz et al., 2024 for a review). The effect of the number of spectral channels in CI-simulated studies indicates that higher number of channels typically provide CI listeners with more distinct spectral information and are accompanied by more accurate phonemes (e.g., Shannon, 1995), words (e.g., Loizou & Dorman, 1999) and sentence recognition (e.g., Gonzalez & Oliver, 2005), specifically in background noise. However, this improvement plateaus after a certain number of channels, with scores remaining unchanged beyond certain points (e.g., Shannon, 1995; Loizou & Dorman, 1999; Gonzalez & Oliver, 2005; Cychosz et al., 2024). The level of channel interaction is typically simulated using varying spectral bandwidths or filter slopes when processing speech signal into continuous spectral bands (e.g., Başkent & Shannon, 2006; Shannon, 1995). Overall, the findings are consistent in showing less overlap between spectral channels in their electrical stimulation provides listeners with more distinct frequency information (e.g., Mehta et al., 2020; Cychosz et al., 2024). However, the filter slope values used to simulate channel interaction in CI-simulated studies may not necessarily replicate the effects of channel interaction on speech perception in actual CI listeners (e.g., Goupell, 2015). Therefore, speech recognition scores from CI-simulated studies may not be directly comparable to outcomes observed in actual CI listeners.

Findings on the effects of simulated insertion depth are mixed and seem to depend, in part, on the listener’s ability to recalibrate spectral perception following the frequency shift introduced by implantation (e.g., Zhou et al., 2010; Geurts & Wouters, 2005). Findings from these simulation studies suggest that frequency-place mismatch is a more detrimental issue than insertion depth alone (Canfarotta et al., 2020) and that modifying this feature does not fully mitigate its effects. Simulation studies provide valuable insights, as they allow researchers to manipulate parameters in controlled environments and isolate specific effects (e.g., Cucis et al., 2021; Xu et al., 2020; DiNino et al., 2016), which would be challenging in studies including CI listeners due to sample size limitations and individual variability. However, the main limitation of simulation studies is that they cannot fully capture all aspects of real-world CI listening. Individual factors, such as age at implantation and the duration of auditory deprivation, are not accounted for in these simulations. Furthermore, evidence suggests that CI listeners differ from normal-hearing individuals in their cortical processing of speech, with reduced activity in the superior and middle temporal gyri (see Farrar et al., 2024 for a review), an aspect not well represented in simulation studies. Given these strengths and limitations, the current study discusses findings from actual CI listeners to provide clinically relevant conclusions while recognizing the complementary role of simulations in advancing our understanding of CI speech processing.

## 2. Methods

### 2.1. Eligibility Criteria

We included all studies published by January 2025. Although this is a narrative review, we employed a structured and transparent approach to the selection and evaluation of studies to synthesize findings relevant to our research questions. We adopted strategies commonly used in systematic reviews to enhance the rigor of our process. The search process is explained under the study selection section. A PIO framework was adopted for this review, where P (patient) referred to postlingually deafened adult CI listeners, I (intervention) was the direct assessment measure(s) for spectral resolution ability based on psychophysical approaches, and O (outcome) was the speech perception score(s). For this review, spectral resolution assessment is considered an intervention within our PIO framework, which is used to structure our research question and conduct an evidence-based analysis. Only studies that assessed spectral resolution based on psychophysical approaches were included while studies using physiological and neurophysiological approaches were excluded. Psychophysical methods are perceptual approaches in which participants directly respond to auditory stimuli with varying frequency content in behavioral tasks.

### 2.2. Information Sources and Search

Six electronic databases were searched to identify relevant publications, namely Academic Search Complete, Google Scholar, MEDLINE (ProQuest), PubMed, Scopus, and Taylor & Francis. These databases were accessed through the University of South Carolina’s library website. Key search terms were identified based on the main goal of this study. Synonyms and abbreviations of the identified key search terms were used to ensure that all relevant publications were included. These five databases were consistently searched using the following combination of key search terms: “*speech perception*” OR “*speech*” OR “*speech recognition*” OR “*word recognition*” OR “*vowel identification/recognition*” OR “*consonant identification/recognition*” OR “*sentence recognition*” OR the same key search terms combined with “*in quiet*” or “*in noise*”, AND “*adult cochlear implant listeners/users/recipients*” OR “*postlingually deafened adult cochlear implant listeners/users/recipients*” AND “*spectral resolution*” OR “*frequency selectivity*”.

### 2.3. Study Selection

This initial search identified 111 papers. The study selection process began with a title and abstract screening, during which duplicate studies were removed. Studies were included if they met the following criteria: (1) participants were postlingually deafened adults with CI, (2) spectral resolution ability was assessed using a psychophysical method, (3) a speech recognition evaluation was conducted, (4) the relationship between spectral resolution and speech perception was statistically tested, and (5) the study was published in English. Exclusion criteria included studies where (1) participants were not actual CI listeners (e.g., studies using vocoder simulations in TH listeners), (2) the focus was on temporal resolution not spectral resolution, (3) study population was children, and (4) the study was a review paper rather than an original research article. After applying these criteria, twenty-four papers remained for full-text review. Busby (1993) was excluded because it did not statistically analyze the relationship between spectral resolution and speech perception. Additionally, papers that were solely focused on method and test development are not included in the table; however, their technical aspects are discussed (e.g., Landsberger et al., 2019; Aronoff & Landsberger, 2013). Among these studies, some also included a control group of participants with typical hearing or hearing aid users; however, this review focuses exclusively on their findings from CI listeners. Table 1 describes the key characteristics and findings of each study.

Table 1. Summary of the findings from prior studies investigating the relationship between measures of spectral resolution ability and speech recognition outcomes in postlingually deafened adult CI listeners. CI: cochlear implant, TH: typical hearing. yo: years old, *R^2^*: coefficient of determination, SNR: signal-to-noise ratio.

## 3. Results

### 3.1. Participants Characteristics

The average sample size across studies was 42 participants per study (SD = ± 93.86). However, after excluding one study with a relatively large sample size of 477 (Gifford et al., 2018), the average is 21 participants per study (SD= ±13.4), with sample sizes ranging from 8 to 31 participants. This shows that the number of participants varied largely across studies, which may influence the statistical power of studies and the robustness of their conclusions. The mean age at implantation across these studies ranged from 47 to 62.5 years old with an average duration of CI experience ranging from 9.78 months to 9.69 years. In terms of the participants’ hearing configuration (unilateral versus bilateral versus bimodal), five studies tested only unilateral CI listeners (Arjmandi et al., 2025; Holden et al., 2016; Gifford et al., 2018; DeVries & Arenberg 2018a; Won et al., 2015) and six studies involved bilateral or bimodal listeners as well (Won et al., 2007; Hughes & Stille 2008; Gifford et al., 2014; Winn et al., 2016; Lawler et al., 2017; DiNino et al., 2020). One study included only bilateral CI listeners (Winn & Litovsky 2015), and the remainder of the reviewed papers did not clearly specify participants’ listening configuration.

### 3.2. Key Findings

Nineteen studies reported a statistically significant association between their measures of spectral resolution and speech recognition test scores (Arjmandi et al., 2025; DiNino et al., 2020; Gifford et al., 2018; Lawler et al., 2017; Hoden et al., 2016; Winn et al., 2016; Winn & Litovsky, 2015; Won et al., 2015; Jeon et al., 2015; Drennan et al., 2014; Gifford et al., 2014; Anderson et al., 2012; Anderson et al., 2011; Saoji et al., 2009; Litvak et al., 2007; Won et al., 2007; Henry et al., 2005; Henry & Turner, 2003; Donaldson & Nelson, 2000; Nelson et al., 1995). Various methods used to assess spectral resolution have demonstrated significant association with speech perception outcomes. These measures include *spectral ripple discrimination threshold* (SRD; Winn et al., 2016; Jeon et al., 2015; Anderson et al., 2011; Won et al., 2007; Henry & Turner, 2003) and its specific modifications for spectral or temporal modulations (DiNino et al., 2020; Lawler et al., 2017; Won et al., 2015; Litvak et al., 2007; Henry et al., 2005). These modified versions of SRD include spectral peak resolution threshold (Henry et al., 2005), clinical SRD (Drennan et al., 2014), *spectral-temporally modulated ripple test* (SMRT; DiNino et al., 2020; Lawler et al., 2017; Holden et al., 2016), two subcategories of SRD called *spectrotemporal modulation detection threshold* (STM; Won et al., 2015), *spectral modulation detection threshold* (SMD or SMT; Litvak et al., 2007; Saoji et al., 2009), as well as a quick version of SMD (QSMD) (Gifford et al., 2014; Gifford et al., 2018). Among these approaches, SRD was the most used approach for evaluating spectral resolution (Winn et al., 2016; Jeon et al., 2015; Anderson et al., 2011; Won et al., 2007; Henry & Turner, 2003). Electrode pitch ranking also showed a moderate significant correlation with speech perception in two earlier studies (Zwolan et al., 1997; Nelson et al., 1995).

Speech perception in the reviewed studies was assessed using recognition of sentences (Gifford et al., 2018; Holden et al., 2016), key words in sentences (Lawler et al., 2017; Won et al., 2015; Anderson et al., 2011; Anderson et al., 2012; Nelson et al., 2011; Zwolan et al., 1997), monosyllabic (Gifford et al., 2018; DeVries & Arenberg, 2018a; Winn et al., 2016; Holden et al., 2016; Winn & Litovsky, 2015; Drennan et al., 2014; Gifford et al., 2014; Anderson et al., 2011; Anderson et al., 2012; Hughes & Stille, 2009; Won et al., 2007; Zwolan et al., 1997) or disyllabic words (Jeon et al., 2015; Won et al., 2007), and phonemes (Arjmandi et al., 2025; DiNino et al., 2020; Jeon et al., 2015 ; Anderson et al., 2012; Anderson et al., 2011; Nelson et al., 2011; Hughes & Stille, 2008; Litvak et al., 2007; Henry et al., 2005 ;Henry & Turner, 2003; Donaldson & Nelson, 2000; Zwolan et al., 1997; Nelson et al., 1995). These assessments were conducted either in quiet conditions (Gifford et al., 2018; DeVries & Arenberg, 2018a; Winn et al., 2016; Holden et al., 2016; Won et al., 2015; Jeon et al., 2015; Anderson et al., 2011; Anderson et al., 2012; Nelson et al., 2011; Hughes & Stille, 2009; Won et al., 2007; Henry et al., 2005; Henry & Turner, 2003) or in the presence of various types of background noise, including multi-talker babble (Gifford et al., 2018; Lawyer et al., 2017; Holden et al., 2016; Won et al., 2015; Jeon et al., 2015; Anderson et al., 2011; Anderson et al., 2012), steady-state noise (Won et al., 2015; Won et al., 2007), and speech-shaped noise (Nelson et al., 2011).

Studies using SRD have found that it accounts for 25% to 59% of the variance in sentences (Anderson et al., 2011), words (Won et al., 2007; Winn et al., 2016; Jeon et al., 2015; Drennan et al., 2014), or phoneme recognition scores (Jeon et al., 2015; Anderson et al., 2011; Henry & Turner, 2003), across both quiet and noisy listening conditions, as indicted by the reported *R^2^* values (Jeon et al., 2015; Anderson et al., 2011; Won et al., 2007). However, some studies did not evaluate speech perception in both listening conditions (Winn et al., 2016; Henry & Turner, 2003), or the significant relationship was observed only in quiet or noise (Anderson et al., 2011). Four studies focused on vowel identification in /hVd/ context (Henry & Turner 2003; Dinino et al., 2020) and CNC word recognition only in quiet (Drennan et al., 2014; Winn et al., 2016), which found a significant relationship between SRD and speech recognition score. One study using SRD found no significant correlation with vowel identification in either quiet or noise, nor with any other speech materials in noise, except for word recognition in sentences presented in quiet (Anderson et al., 2011). However, in a subsequent study, Anderson et al., (2012) reported strong significant correlations in both quiet and noisy listening conditions when using spectral ripple detection thresholds, also referred to as the spectral modulation transfer function (SMTF). This finding suggests that SMTF may be a more effective measure compared to SRD for establishing the relationship between spectral resolution and speech perception. It also emphasizes the distinction between two tasks of spectral ripple discrimination versus detection threshold; however, previous studies have not directly compared them within the same study to indicate how similarly or differently they predict speech perception outcomes (Anderson et al., 2012; Gifford et al., 2018; Saoji et al., 2009; Litvak et al., 2007) and the discrimination task is more widely used. Overall, the findings from using SRD indicate that this measure is a relatively reliable measure of spectral resolution when examining the association between the ability to resolve spectral information and speech recognition scores in postlingually deafened adult CI listeners in both quiet and noisy conditions.

Furthermore, although performance on this test may be influenced by how individual electrodes contribute to spectral information transmission, it is a global measure of spectral resolution rather than a channel-specific measure. As such, it does not provide detailed information about the performance of individual CI electrodes, which may limit its utility for identifying specific cochlear regions with poor spectral resolution. While this limits its ability for targeted programming modifications (e.g., channel deactivation or current focusing), it remains valuable for evaluating the overall effects of CI programming strategies and auditory training on global spectral resolution. It is worth noting that SRD captures the amplitude modulation aspects of speech perception in CI listeners; however, it does not provide a pure assessment of spectral resolution ability given the possible confounding factors that will be discussed in the following section (Aronoff & Landsberger, 2013). Instead, the *spectral-temporally modulated ripple test* (SMRT) introduces a modified version of SRD and a more precise method of assessing spectral resolution. Lawler et al. (2017) utilized SMRT and the Arizona Biomedical Institute (AzBio) sentence test in noise at an 8 dB signal-to-noise ratio (SNR). The same test materials besides CNC word recognition in quiet and Hearing in Noise Test (HINT) of sentence recognition were utilized in Holden et al., (2016). In addition, one study examined this relationship with vowel identification in quiet (DiNino et al., 2020). All three studies found a moderate significant association between SMRT and their test of speech perception with *R^2^* values ranging from 0.33 to 0.69. These findings suggest that SMRT is a sensitive measure for assessing the relationship between spectral resolution and speech perception across different types of speech stimuli.

Studies that used SMT or SMD test to evaluate spectral resolution ability reported significant correlations with phoneme recognition in quiet (*R²* = 0.7 for vowel identification and *R²* = 0.67 for consonant identification; Litvak et al., 2007; Saoji et al., 2009). A modified, clinically-validated version of the test, QSMD, has also shown significant relationship with speech recognition scores. Gifford et al. (2014) reported an *R*² of 0.66 between QSMD scores and CNC word recognition in quiet, whereas Gifford et al. (2018) observed a substantially lower *R*² of 0.27 for the same measure. Additionally, AzBio sentence recognition in quiet and in noise at a 5 dB SNR was correlated with QSMD scores (*R*² = 0.26; Gifford et al., 2018). Although the QSMD addresses the limitation of the original SMD test in being time-consuming, the marked variability in *R*² values is noteworthy, especially given the similarities in participant demographics and test materials across studies (see Table 1). In particular, the significant drop in *R*² in the larger sample study (*N* = 477) compared to the smaller sample (*N*=76) suggests that increased variability across participants may be associated with attenuation in the observed relationships.

Electrode pitch ranking is another method used to assess spectral resolution in CI listeners. It accounted for a moderate portion of the variance in consonant recognition scores in quiet (Donaldson & Nelson 2000; Nelson et al., 1995), particularly when the relationship was examined using the relative transmitted information specific to place of articulation (Donaldson & Nelson 2000). It should be noted that the perceptual separation of electrodes, rather than their fixed physical distance along the array, played a role in listeners’ ability to rank pitch. A distinct pitch sensitivity and significant improvement in consonant recognition was achieved only at certain spatial distance between electrodes. In Donaldson & Nelson (2000), the average distance was above 4.5 mm, while Nelson et al. (1995) reported a wide range, from 0.47 mm to 8.71 mm. Furthermore, in Donaldson & Nelson (2000), the significant correlation between pitch sensitivity and consonant recognition was observed only among participants who were experienced users of the spectral peak (SPEAK) speech processing strategy with at least one year of CI use. This strategy was designed to provide more detailed spectral characterization of the speech waveform (Skinner et al., 1994). This significant association was not, however, observed in the Multipeak Speech Processor (MPEAK) users who had recently switched to SPEAK, which emphasizes the importance of both programming strategy and duration of hearing through CI.

On the other hand, a group of studies reported no statistically significant relationship between their measures of spectral resolution and speech perception. Forward masking psychophysical tuning curve (PTC) or forward masking spatial tuning curve (fmSTC), and the electrode discrimination test have generally not shown significant correlations with phoneme, word, and sentence recognition scores in adult CI listeners (DeVries & Arenberg 2018a; Anderson et. al., 2011; Nelson et al., 2011; Hughes & Stille 2009; Zwolan et al., 1997). A recent study found a modest but statistically significant correlation between PTCs and vowel identification in noise (*R²* = 0.21; Arjmandi et al., 2025), suggesting that PTCs may be more predictive of speech perception under complex listening conditions. Notably, this study quantified spectral resolution using the normalized slope of the tuning curve, rather than peak threshold, which may account for the observed effect. Although these approaches provide region-specific estimates of spectral resolution, further research is needed to elaborate the inconsistent findings and clarify their relationship to speech perception outcomes.

Overall, these findings demonstrate that the relationship between spectral resolution and speech perception appears to be dependent on the specific measurement approach. While some approaches, such as SRD, have consistently demonstrated a robust correlation with speech outcomes (Winn et al., 2016; Jeon et al., 2015; Anderson et al., 2011 & 2012; Won et al., 2007; Henry & Turner, 2003), others, including PTC, have yielded more variable and often inconclusive results (DeVries & Arenberg 2018a; Anderson et al., 2011; Nelson et al., 2011; Hughes & Stille 2009). Even among the more successful approaches, reported R² values vary widely, reflecting differences in predictive power and practical utility, likely due to methodological variations, characteristics of the study populations, and the multifactorial nature of speech perception itself. Beyond predictive accuracy, it is important to evaluate these measures based on some additional practical criteria. Time efficiency is particularly important, as assessments that are overly time-consuming or resource-intensive may be impractical for routine clinical use. Another key consideration is whether a measure provides a global estimate of spectral resolution or offers region-specific information along the cochlea. Global measures, while useful for evaluating overall performance (e.g., both before and after changes to CI mapping parameters, as well as in the context of auditory training), do not capture the spatial variability in auditory function across the cochlea. Several studies have shown that ENI quality, channel interaction, and frequency selectivity vary across different regions of the cochlea (e.g., Arjmandi et al., 2022a; Jahn et al., 2020; Abbas et al., 2004). Therefore, methods capable of assessing spectral resolution at the level of specific cochlear regions or groups of electrodes may support more targeted programming strategies, such as the deactivation of electrodes associated with poor ENI quality.

Arguably, a test of spectral resolution should be evaluated based on its ability to capture the categorization nature of speech perception beyond pure sound discrimination (Holt & Lotto, 2010; Winn & Litovsky, 2015). Speech perception involves interpreting continuous acoustic signals and mapping them onto distinct linguistic categories, such as phonemes or words. This categorization process relies on access to both coarse- and fine-grained acoustic information and the ability to group similar sounds into meaningful units (Holt & Lotto, 2006; 2010). Traditional measures of spectral resolution, however, often focus on the ability to distinguish between isolated frequencies or tones and may not fully capture how listeners categorize complex, speech-like signals. As a result, these measures might overlook important aspects of how the spectral resolution ability of CI listeners relates to their ability to process, perceive, and understand speech. At the same time, tone-based or non-speech spectral modulation stimuli offer the advantage of minimizing the confounding effects of higher-level linguistic processing, which, although not yet empirically demonstrated, might confound or inflate performance on speech-based tasks. Finally, an effective measure should account for the unique nature of signal transmission in CIs, which conveys modulation frequency within and across spectral bands (e.g., Zeng et al., 2008; Goupell, 2015). This distinction may be important depending on the intended use of the measure, as measures that do not align with how CIs encode spectral cues might only partially reflect the relationship between spectral resolution and speech perception outcomes.

Taken together, these considerations emphasize the need for selecting or developing measures that balance predictive accuracy with clinical practicality. The observed variability across results may indicate the influence of several factors, including the choice of speech stimuli (sentence or words or phonemes), noise type (static or multi-talker babbling or speech-shaped noises), specific features of spectral resolution tests such as ripple density in SRD, as well as individual differences in cortical processing of spectral information in speech (Bálint et al., 2024; Farrar et al., 2024; Finke et al., 2016). The latter remains understudied, and there is limited information available on cortical processes in CI listeners in response to variations in spectral resolution and speech-evoked cortical activities (see Farrar et al., 2024 for a review).

### 3.3. Synthesis of Results

Several factors may contribute to the variable findings observed across these studies. These factors can be broadly categorized into the assessment methods used to measure spectral resolution and those used to evaluate speech perception. Figure 1 provides a visual summary of the patterns observed in the statistically significant findings. In the following sections, we will discuss these factors in detail and compare findings to better understand their potential influences on the relationship between spectral resolution and speech perception in adult CI listeners.

**Figure 1.**
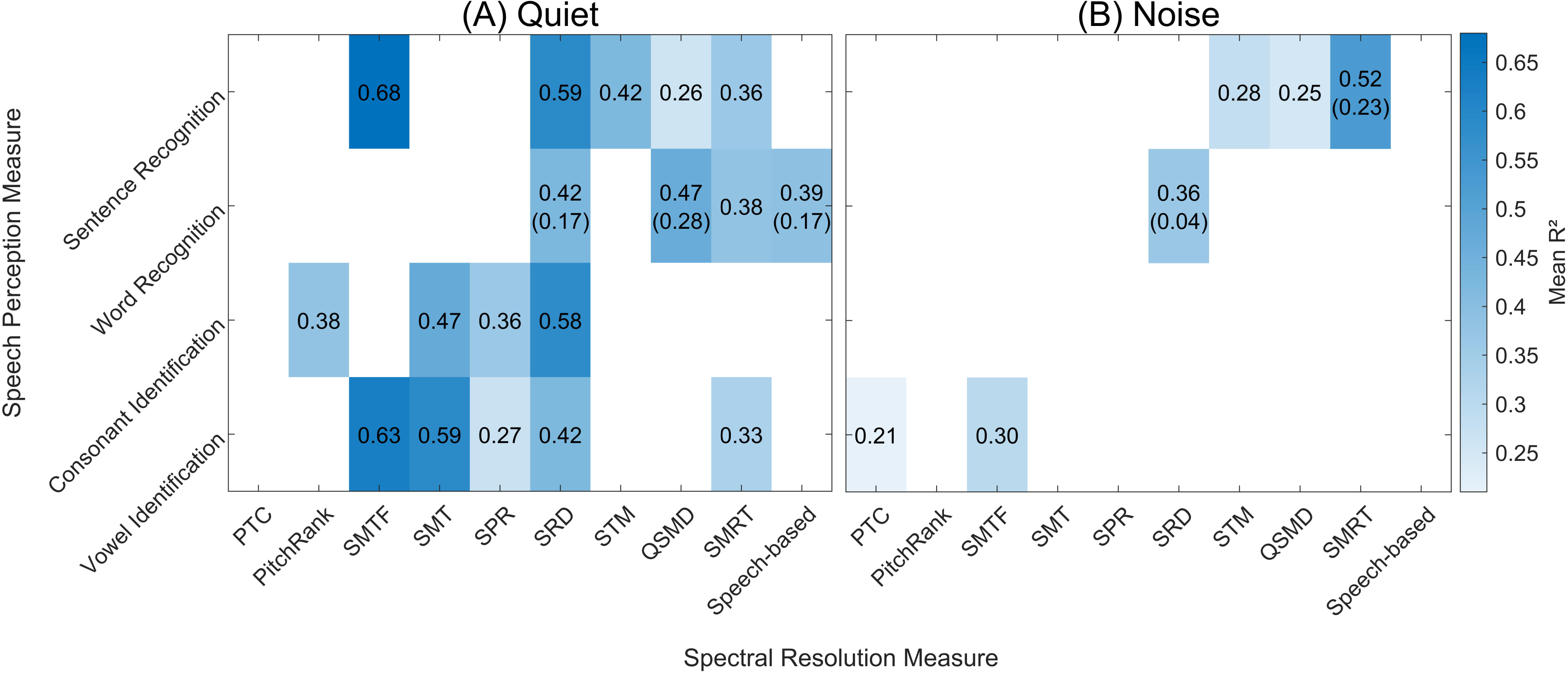
Heatmap showing *R²* values for associations between spectral resolution (x-axis) and speech perception measures (y-axis) in quiet (A) and noise (B). To ensure meaningful interpretation, only studies that reported statistically significant associations (*p* < .05) are included. Each cell displays the mean *R²* value, with standard deviation in parentheses; cells without SD reflect a single study. Color intensity represents the strength of the association, with light blue indicating lower *R²* values and darker blue indicating stronger associations. A color bar on the right provides the *R²* scale. PTC: Psychophysical tuning curve, PitchRank: Electrode pitch ranking, SMTF: Spectral modulation transfer function, SMT: Spectral modulation threshold, SPR: Spectral peak resolution, SRD: Spectral ripple discrimination threshold, STM: Spectrotemporal modulation detection, QSMD: Quick spectral modulation detection, SMRT: Spectral-temporally modulated ripple test.

#### 3.3.1. Potential Impacts of Spectral Resolution Measures

There are fundamental differences in the methodologies used to evaluate spectral resolution in the reviewed studies. The *R^2^* values suggest that some of these approaches are more reliable in exploring the relationship between spectral resolution ability and speech perception. The SRD test has been extensively used for this purpose (Winn et al., 2016; Jeon et al., 2015; Anderson et al., 2011 & 2012; Won et al., 2007; Henry & Turner, 2003). In this test, the listener is presented with broadband noises with amplitude-modulated (rippled) power spectra (e.g., Supin et al., 1994; Supin et al., 1997; Supin et al., 1999). The task is to differentiate between a reference and a target sound, where the target has its spectral peaks and valleys inverted in a phase compared to the reference sound. As the definition implies, this test includes three manipulable components: *modulation depth*, *modulation rate* (frequency), and *ripple density* (frequency of ripples per octave, RPO). Modulation depth and rate are used to assess individuals’ access to temporal information in the signal envelope, while ripple density involves a spectral manipulation applied to the envelope of the stimuli. Discrimination of a smaller modulation depth, higher temporal modulation rate, and a larger number of ripples per octave is typically interpreted as a sign of better spectral resolution ability (e.g., Winn et al., 2022; Winn et al., 2016). Many of these studies used a typical spectral ripple discrimination task, which is conducted using a modulation depth of 30 dB and temporal modulation rate of 5 or 10 Hz with adaptive ripple frequency or density starting from 0.5 ripple per octave (Lawler et al., 2017; Won et al., 2015; Jeon et al., 2015; Drennan et al., 2014; Anderson et al., 2011; Henry & Turner 2003).

To examine the effect of temporal or spectral modulation of signal on spectral resolution results, each of these features can be manipulated while keeping the other features constant. To investigate the effect of spectral ripple frequency, some studies looked at a number of densities including 0.5, 1 and 2 RPO, which are considered low densities (Winn et al., 2016; Won et al., 2015; Litvak et al., 2007). Another study evaluated these low densities with more detail at 0.25, 0.75 and 1.5 RPO in addition to the 0.5, 1 and 2 RPO densities (Anderson et al. 2012). To investigate the effect of higher ripple densities, some studies explored a wide range of ripple frequencies per octave with 13 (Henry & Turner, 2003) or 14 different RPOs, including 0.125, 0.176, 0.25, 0.354, 0.5, 0.707, 1.0, 1.414, 2.0, 2.828, 4.0, 5.657, 8.0, and 11.314 RPO (Jeon et al., 2015; Drennan et al., 2014; Won et al., 2007; Henry et al., 2005). While a general finding from these studies show a significant correlation exists between ripple discrimination threshold task (ripple detection in Anderson et al., 2012) with speech perception, it is also evident that high ripple densities result in some level of aliasing (Anderson et al., 2011; Winn & O’Brien, 2021) and temporal smearing of the amplitude modulation (Lawler et al., 2017), particularly in CI listeners. Also, some of the observed relationships between our target variables, spectral resolution and speech perception, were only evident with low ripple densities (Won et al., 2015; Anderson et al., 2012). Other studies used QSMD as a faster version of SRD that requires the listener to discriminate a flat spectrum noise from one with spectral peaks and valleys. In this design, target signal is presented with five modulation depths (10, 11, 13, 14, and 16 dB) and two modulation frequencies (0.5 and 1.0 cycle/oct) (Gifford et al., 2014 & 2018). They found significant but variable correlations with CNC word recognition and AzBio sentence recognition in quiet and with multi-talker babble noise in a +5 dB SNR (Figure 1). Drennan et al., (2014) modified the conventional SRD test with randomized spectral ripple envelope phase, 12 ripple densities (0.125, 0.176, 0.25, 0.354, 0.5, 0.707, 1.0, 1.414, 2.0, 2.828, 4.0, 5.657 RPO) and denser noise, introduced as clinical SRD. After being validated and showing high correlation with the conventional approach, clinical SRD successfully explained an average of 58% variation in CNC word recognition scores in 28 postlingually deafened adult CI listeners (Drennan et al. 2014).

As highlighted by Azadpour and McKay (2012), several factors could complicate the interpretation of SRD test results. One such factor is the emergence of local loudness cues. For example, if a listener concentrates on a narrow frequency band that does not cover a complete ripple cycle, any inherent loudness difference between the target and reference stimuli in that band may serve as an unintended cue. Additionally, cues might arise from the extreme frequency edges of the stimuli. When listeners focus on either the high or low frequency limits, differences in the maximum or minimum audible frequencies between the two stimuli could inadvertently signal the distinction. Moreover, the overall balance of frequencies, quantified as the spectral centroid or weighted mean frequency, might shift between stimuli. If listeners are sensitive to this shift, they may perceive an audible difference even when the primary manipulation is intended to affect only the ripple structure. Collectively, such confounding factors can lead to overestimation of spectral resolution measurement via SRD in CI listeners (Aronoff & Landsberger, 2013; Winn & O’Brien, 2022). SMRT is designed to eliminate the role of these extraneous cues in spectral resolution by drifting the modulation phase over time in both target and reference stimuli (Aronoff & Landsberger, 2013). This test has shown significant association with vowel identification in quiet (Dinino et al., 2020) and with AzBio sentence recognition in noise (Holden et al., 2016; Lawler et al., 2017), as well as sentence and word recognition in quiet (Holden et al., 2016). To account for these confounding factors in SRD, Won et al. (2015) also used a modified version of SRD, STM, and showed significant association with sentence recognition scores even after controlling for the effect of temporal modulation detection. Notably, the *R^2^*values for STM and sentence recognition slightly increased after accounting for this effect (from 0.38 to 0.42 for K-CID and from 0.22 to 0.28 for K-HINT), though this increase may not be statistically meaningful.

Another psychophysical approach used for the purpose of spectral resolution assessment is forward-masking PTCs, which involves presenting a brief probe stimulus at a constant, low current level (10%–30% of the dynamic range), which is expected to activate a small region of neural elements associated with the test electrode (i.e., probe electrode; Nelson, & Freyman, 1984; Shannon, 1990). Forward masking is used to determine the masker level at which the masker signal barely masks the probe signal. The masker signal can vary in location along the electrode array, ranging from the probe electrode (on-frequency masker) to electrodes positioned on either apical or basal sides relative to the probe electrode. The forward-masking spatial tuning curve (fmSTC) or PTC is a graph that illustrates the masker level on different electrodes that just mask a probe stimulus fixed at a specific level (Moore, 1978; Arenberg et al., 2022; Arjmandi et al., 2025). In other words, it shows how the level of a masking stimulus, presented shortly before the probe, must vary across electrodes to effectively mask the perception of the probe stimulus as a separate tone. Typically, masker levels are lowest near the probe electrode and increase with distance from the probe electrode, both apically and basally, resulting in a tuning curve shape (i.e., PTC). The sharpness of PTC is typically quantified as an indicator of spectral resolution on the probe electrode, with sharper PTCs indicating greater spectral resolution for the location of probe electrode along the electrode array (e.g., Nelson et al., 2008; Nelson et al., 2011). Despite the precise information that PTCs provide about the frequency selectivity of specific cochlear regions stimulated by individual electrodes, prior studies have often reported non-significant correlations between PTC or STC sharpness and speech perception measures (DeVries & Arenberg 2018a; Hughes & Stille 2009; Nelson et al., 2011; Hughes & Stille 2009). One reason for this may be methodological variability across studies, including differences in how PTC sharpness is quantified, which electrodes are tested, and what speech perception measures are used. Additionally, studies often used broad speech outcomes (i.e., sentence recognition) that may not be sensitive enough to reflect deficits in spectral resolution. In contrast, recent findings showed a significant relationship between the mean PTC slope from a mid-array electrode and vowel identification in noise (Arjmandi et al., 2025), a task that places greater demands on access to fine spectral information. This finding suggests that PTC, combined with speech perception tasks that emphasize spectral cues and specific quantification methods, can offer some insights into region-specific spectral resolution and its relationship to speech recognition scores.

Some of the earlier methods for assessing spectral resolution focused on individuals’ sensitivity to place-pitch, that is, the psychophysical perception of the place of stimulation within the cochlea, which serves as a key cue for pitch perception (e.g., Donaldson & Nelson, 2000; Nelson et al., 1995). This approach, which is called electrode pitch ranking, involves asking participants to rank and judge the tones’ frequency presented in a pair of electrodes within certain spatial distances of 0.75, 1.5, 3.0, and 4.5 mm. In some subjects who required more electrode separation, they were tested with extended distances of 6.0 and 7.5 mm (Donaldson & Nelson, 2000). Accurately ranking two closely spaced electrodes indicates good ENI quality and spectral resolution ability (e.g., Townshend et al., 1987; Nelson et al., 1995). This approach was specifically tested for its association with consonant identification, with a focus on analyzing how well place of articulation information is transmitted (Donaldson & Nelson, 2000; Zwolan et al.1997; Nelson et al. 1995). This specific cue in consonant identification requires precise access to spectral information (e.g., Sydral, 1983). The analysis was centered on groups of consonants where differentiation relies heavily on their place of articulation, such as /m/ versus /n/ sounds or /f/ versus /v/ sounds. These studies showed considerable variability among CI listeners in place-pitch sensitivity (e.g., Donalson & Nelson, 2000). However, a general trend observed was that CI listeners with better place-pitch sensitivity, measured as accurate pitch ranking in closely spaced electrodes, tended to have better consonant recognition due to improved utilization of spectral cues (Donaldson & Nelson 2000; Nelson et al., 1995). It was found that participants needed varying amount of spatial separation between stimulation points on the cochlea, ranging from 0.47 mm to 8.71 mm, to achieve a certain level of sensitivity to place-pitch differences. This degree of spatial separation explained 37% of the variability in how accurately participants perceived the place-of-articulation features of consonants (Donaldson & Nelson 2000). This significant correlation was achieved only with spatial separation above 4.5 mm in one group (Nelson et al., 1995).

Another method for evaluating place-pitch sensitivity involved including perceptually discriminable electrodes in individual’s everyday CI map (Zwolan et al., 1997). This method uses an adaptive electrode discrimination task, where participants are asked to determine whether two intervals with different frequencies are perceived as the same or different. Discriminable electrodes were included in an experimental map and a wide range of speech perception tasks were assessed (phoneme, word, and sentence recognition). The results varied widely among the participants, but a subgroup of participants showed a significant increase in their speech perception scores when using only the discriminable electrodes in their CI programming. This finding demonstrates the value of considering electrode-specific tasks when measuring the relationship between spectral resolution and speech perception. However, even when using region-specific tasks to assess spectral resolution, the ability to predict speech perception may vary depending on the type of speech stimuli and individual differences among participants.

Finally, in an attempt to capture the nature of speech perception as a categorization task using speech cues, studies have proposed using speech-based tasks that involve categorization of speech sounds that heavily rely on resolving spectral information (Winn et al., 2016; Winn & Litovsky, 2015; Dinino et al., 2020). These tasks require access to multiple spectral cues. For vowel categorization, listeners rely on formant transitions (e.g., Donaldson et al., 2015), which reflect rapid changes in the resonant frequencies of the vocal tract and serve as a fine spectral cue, and spectral tilt (e.g., Rødvik et al., 2018), which provides information about the overall shape of the spectrum and serves as a coarse spectral resolution cue. For consonant categorization, particularly in stop and fricative contrasts, listeners depend on place-of-articulation cues, which are conveyed by spectral patterns such as the frequency and bandwidth of noise bursts or formant transitions (e.g., Rødvik et al., 2018), requiring both fine and coarse spectral resolution. Studies have shown that the ability of CI listeners to use fine spectral information, such as formant transitions, is correlated with their ability in performing various speech perception tasks. For example, Winn & Litovsky (2015) and Winn et al. (2016) found correlations between formant-based spectral resolution tasks and word recognition scores. Similarly, DiNino et al. (2020) linked formant cue sensitivity to vowel identification. These findings indicate that speech-based assessments of spectral resolution may provide a promising approach for investigating the relationship between spectral resolution ability and speech perception in an ecologically valid manner.

Current evidence supporting the effectiveness and generalizability of this approach remains limited (Figure 1). To date, studies have primarily utilized a narrow set of speech stimuli, such as /ba/ versus /da/ pairs (Winn et al., 2016; Winn and Litovsky, 2015; DiNino et al., 2020), which do not encompass the broader range of frequency information transmitted through CI electrodes for different vowel and consonant sounds. Also, the strength of this approach in capturing the relationship with speech perception dropped noticeably from one study (*R*^2^ = 0.5, Winn et al., 2016) to another (*R*^2^ = 0.27, Winn & Litovsky, 2015). Additionally, consonant categorization using spectral tilt cue failed to show a relationship with either word or vowel recognition. Investigating a more comprehensive approach with a larger set of speech stimuli may help address these limitations, allowing for a more complete characterization of variations in spectral resolution across cochlear regions, as well as their impact on different levels of speech perception.

#### 3.3.2. Potential Impacts of Speech Perceptions Measures

The specific type of speech test being used may affect the ability to perceive and understand speech. Speech perception is a task-dependent ability, and the context essentially affects speech perception scores. Although often correlated, speech recognition scores may vary depending on whether individuals are asked to identify vowels, consonants, monosyllabic words, key words in sentences, or entire sentences. Therefore, observed differences in the relationship between measures of spectral resolution and speech perception across studies may be partly due to the variations in the speech materials used. We did not conduct a statistical analysis to determine how variations in speech perception measures contribute to discrepancies in linking spectral resolution and speech perception, as such an analysis would be limited by cross-study differences in participant characteristics (e.g., demographic, clinical, and cognitive abilities) that confound interpretation. Instead, we provide an overview of the types of speech stimuli used across the reviewed studies.

Tests of sentence recognition included AzBio, Central Institute for the Deaf (CID), HINT, City University of New York (CUNY), Institute of Electrical and Electronics Engineering (IEEE), and Bamford-Kowal-Bench (BKB) standard sentences. In nearly all sentence recognition tasks, the percentage of correct responses was determined based on the number of accurately identified key words. For word recognition tasks, monosyllabic CNC word lists from Peterson & Lehiste, (1962), Northwestern University Auditory Test Number Six (NU-6), and Maryland University were the most used. Additionally, two studies assessed word recognition ability using spondee word lists (Won et al., 2007; Jeon et al., 2015). Phoneme recognition was also used for this purpose in which participants were tested for vowel recognition in h-vowel-d (/hVd/) contexts (DiNino et al., 2020; Anderson et al., 2011; Anderson et al., 2012; Nelson et al., 2011; Henry et al., 2005; Henry & Turner, 2003; Zwolan et al., 1997) or b-vowel-t (/bVt/) contexts in one study (Litvak et al., 2007) and consonant identification within an a-consonant-a (/aCa/) context (Jeon et al., 2015; Nelson et al., 2011; Litvak et al., 2007; Henry et al., 2005; Donaldson & Nelson, 2000; Zwolan et al., 1997; Nelson et al., 1995). The percentage of correct phonemes was also evaluated during word recognition in some of these studies (Hughes & Stille, 2009; Zwolan et al., 1997). In speech-based measures, three studies manipulated spectral cues in six consonant vowel syllables, which included liquids */ra/* versus */la/*, fricatives */sa/* versus */ a/*, and plosives */ba/*-*/da/*, but their group analysis was restricted to responses to syllables in the */ba/-/da/* continuum (Winn et al., 2016; Winn & Litovsky 2015; DiNino et al., 2020).

Arguably, conventional speech perception measures are not sensitive enough to capture spectral resolution ability. Speech is inherently a multidimensional stimulus, with multiple cues working together to shape perception abilities (e.g., Toscano & McMurray, 2010; Holt & Lotto, 2006; Arjmandi et al., 2018; Rosenblum, 2008; Christensen & Humes, 1997). Considering both temporal and spectral cues involved in speech perception, even when a target sound is accurately identified, it does not guarantee that spectral cues were fully resolved. Listeners with CI may use compensatory strategies in the temporal domain to achieve good speech identification performance despite limited access to spectral cues (e.g., Moberly et al., 2014; Fleming & Winn, 2022; Peng et al., 2012; Winn et al., 2012). Cues such as consonant burst spectrum and formant transitions in consonants or formant frequencies in vowels are reliable representatives of spectral cues (Hillenbrand et al., 1995; Lehiste, 1965). Therefore, vowel identification and consonant categorization tasks that manipulate spectral cues (e.g., the first and second formant frequencies, cues for place of articulation) can reveal how effectively listeners use these spectral cues for speech perception. As an example from CI-simulation studies, DiNino et al. (2016) showed that altering specific aspects of CI channel function, such as deactivating certain channels (creating holes) or making more overlap between channels (shallower filter slope), influence participants’ abilities to correctly identify vowels, and the errors they made were systematic and predictable based on the type of manipulation. This suggests that speech-based measures of spectral resolution have the potential to provide a clinically relevant approach for establishing the relationship between spectral resolution ability and speech perception outcomes.

Further bolstering clinical relevance, some studies have incorporated speech stimuli presented in background noise to evaluate the role of spectral resolution in understanding speech, making their findings more applicable to real-world settings (Won et al., 2007; Nelson et al., 2011; Anderson et al., 2011 & 12; Jeon et al., 2015; Won et al., 2015; Holden et al., 2016; Lawler et al., 2017; Gifford et al., 2018; Arjmandi et al., 2025). The presence of noise highlights the importance of spectral resolution, as noise can either distort the transmitted information or mask it with additional energy (e.g., Harmon, 2021). Compared to speech perception in quiet, understanding speech in noisy environments requires enhanced access to spectral information (e.g., Baer & Moore, 1993); however, as illustrated Figure 1 panel (B), fewer studies have investigated this relationship while presenting speech stimuli in background noise. In the reviewed papers, types of noise included were steady state (Won et al., 2007), multi-talker babble (Won et al., 2007; Arjmandi et al., 2025; Gifford et al., 2018; Jeon et al., 2015), and speech shaped noise (Nelson et al., 2011) with variable SNRs ranging from -6 dB to +21 dB and more focus on a 0 dB to 10 dB SNR (Arjmandi et al., 2025; Gifford et al., 2018; Lawler et al., 2017; Nelson et al., 2011). Studies examining the relationship between spectral resolution and speech perception in both quiet and noisy conditions have found a significant correlation, but the strength of this correlation differed between the two listening conditions (Figure 1). Many of them found a stronger relationship in quiet compared to noise (Anderson et al., 2011; Anderson et al., 2012; Jeon et al., 2015; Won et al., 2015), However, the *R^2^*did not differ substantially in Gifford et al., (2018) and Holden et al., (2016). By contrast, Won et al., (2007) used the steady-state noise for the noisy condition and found stronger correlation between SRD and spondee word recognition compared to CNC word recognition in quiet.

Our review of prior findings on the relationship between spectral resolution and speech perception suggests that the type of speech materials used may influence study outcomes (Henry et al., 2005; Anderson et al., 2011; Winn et al., 2016; Winn & Litovsky 2015; Won et al., 2007). In addition to examples from the effect of listening condition (noise versus quiet), while checking the relationship between the measure of spectral resolution with both vowel and consonant recognition, a stronger relationship was revealed with consonants (Henry et al., 2005). This stronger correlation with consonant recognition was also observed compared to word recognition in noise (Jeon et al., 2015). Additionally, in another study, while a strong correlation was observed between keyword in sentence recognition and SRD, no significant correlation was observed for vowels (Anderson et al., 2011). This could be due to factors such as differing spectral fluctuations among stimuli or varying levels of information redundancy, such as between phonemes compared to sentences (Anderson et al., 2011), which can impact speech perception. However, based on the current findings from the reviewed studies, we cannot attribute non-significant relationships or variability in correlations solely to differences in speech materials.

This conclusion is supported by studies using the same speech tests that have produced varying, or sometimes significantly different, *R²* values in their findings (Henry et al., 2005; Anderson et al., 2011; Winn et al., 2016; Winn & Litovsky 2015; Won et al., 2007). For example, testing vowel identification using the same /hVd/ contexts with SRD demonstrated a relatively strong correlation in Henry et al., (2005; *R^2^*= 0.42), while no significant correlation was observed in Anderson et al., (2011). The strength of observed relationships across studies when used the same speech materials also varied noticeably such as in Winn & Litovsky (2015) and Winn et al., (2016). Some studies employed a comprehensive battery of speech tests to assess spectral resolution and found no correlation between their spectral resolution measures and any of the speech tests, regardless of the specific test characteristics (Nelson et al., 2011; Hughes & Stille 2009; Zwolan et al., 1997). Overall, the findings suggest that differences in spectral resolution assessment methods have a greater impact on capturing the relationship between spectral resolution ability and speech perception than differences in speech perception methods.

## 4. Discussion

We reviewed and synthesized prior findings on the relationship between spectral resolution ability and speech recognition in postlingually deafened adult CI listeners. The primary objective was to discuss studies that employed psychophysical methods to assess spectral resolution and its association with various speech perception tasks. Among the reviewed studies, eighteen (75%) reported a statistically significant relationship between measures of spectral resolution and speech perception, with *R²* values ranging from 0.21 to 0.69 (Arjmandi et al., 2025; DiNino et al., 2020; Gifford et al., 2018; Lawler et al., 2017; Winn et al., 2016; Holden et al., 2016; Winn & Litovsky, 2015; Won et al., 2015; Jeon et al., 2015; Drennan et al., 2014; Gifford et al., 2014; Anderson et al., 2012; Anderson et al., 2011; Litvak et al., 2007; Won et al., 2007; Henry et al., 2005; Henry & Turner, 2003; Donaldson & Nelson, 2000; Nelson et al., 1995). This variation indicates that the strength of the relationship is inconsistent across studies. Some studies found no significant association (Zwolan et al., 1997; Hughes & Stille, 2009; Nelson et al., 2011; DeVries & Arenberg, 2018a). We critically discussed their methodological differences. This evaluation highlights the strengths and limitations of current approaches and their potential for improving our understanding of the link between spectral resolution and speech perception in CI listeners.

### 4.1. Relationship Between Spectral Resolution and Speech Perception

Spectral resolution reflects a listener’s ability to distinguish fine differences in frequency information (Litvak et al., 2007; Won et al., 2007), which is critical for decoding speech sounds, such as distinguishing different vowels and consonants. Speech perception, particularly in noisy environments, relies heavily on spectral resolution ability to access important acoustic cues like formant frequencies, formant transitions, spectral tilt, and place of articulation (e.g., Xu et al., 2005; Rødvik et al., 2018). Accurate understanding of the relationship between spectral resolution ability and speech perception helps pinpoint the mechanisms that contribute to better speech recognition, particularly in the presence of background noise where many CI listeners struggle significantly (e.g., Arjmandi et al., 2022a; Oxenham, & Kreft, 2014; Zeng, 2004; Ashjaei et al., 2022). In addition, spectral resolution varies among individual CI listeners (e.g., Arjmandi et al., 2022a; DeVries & Arenberg, 2018a; Caswell-Midwinter et al., 2022) and even across cochlear regions (Arjmandi et al., 2022a; Arenberg et al., 2018) due to the loss and/or smearing of spectral information caused by channel interaction (e.g., Jones et al., 2013; White et al., 1984; Cucis et al., 2021; Throckmorton & Collins, 2002). Better understanding of this relationship is important for researchers and clinicians to develop reliable tools for effective assessment and targeted interventions, including evaluation of speech processing strategies (e.g., Geurts & Wouters, 2004; Wouters et al., 2015), adjustments to CI programming (e.g., deactivation, current steering, or focusing; Arenberg et al., 2018), and assessment of auditory training approaches (e.g., Moberly et al., 2020).

The observed association between spectral resolution ability and speech perception varied across studies, which could be attributed to differences in the assessment methods, as well as other audiological factors and the demographics of the study population. Among spectral resolution methods, SRD emerges as the most prevalent method for evaluating spectral resolution and accounts for a considerable percentage of the variance in speech recognition scores (25% to 59%). Although SRD shows reliable performance in assessment of spectral resolution and its association with speech recognition, its effectiveness varies depending on the technical details such as ripple density, test condition (quiet versus noise), and participants’ experience with CI (Winn et al., 2016). Modified versions of SRD, particularly SMRT and SMD, also provide insight into this relationship. SMRT could predict on average between 33% to 69% of the variability in speech perception scores, while controlling for the other cues contributing to SRD results. Moreover, while SMD demonstrated significant correlations with phoneme recognition (Litvak et al., 2007; Saoji et al., 2009), its time-consuming nature prompted the development of a faster version, QSMD, which still maintains an association with word recognition (Gifford et al., 2018; Gifford et al., 2014).

The measure of electrode pitch ranking presents an intriguing divergence from region-specific assessments of spectral resolution, as it has shown significant correlations with consonant recognition, particularly when considering the spatial arrangement of electrodes and their influence on pitch perception (Nelson et al., 1995; Donaldson & Nelson, 2000). However, its effectiveness appears highly dependent on background factors such as experience with specific speech processing strategies, as demonstrated by differences in outcomes between SPEAK and MPEAK users (Donaldson & Nelson, 2000). At the same time, several studies have identified methods that lack significant predictive power for speech perception (Zwolan et al., 1997; Hughes & Stille, 2009; Nelson et al., 2011; DeVries & Arenberg, 2018a). For example, PTCs and electrode discrimination tests did not yield consistent correlations with speech perception, highlighting limitations within these assessments. The observed discrepancies across findings underscore the impact of both methodological variations and individual differences among participants. These inconsistencies highlight the need for further refined experimental approaches that better account for the heterogeneous auditory profiles of CI listeners, as well as more controlled studies with carefully selected participant groups to minimize variability and enhance the reliability of findings.

### 4.2. Methodological Insights and Challenges

Observed variabilities in predictive capacity of different measures of spectral resolution can be partly explained by speech materials used in these studies. Sentences, words, and phonemes differ markedly in the level of redundancy and spectral information they provide to listeners (Lorenzi et al., 2006), which can influence their perceptual demands and the cues needed for accurate recognition (Xu et al., 2005; Repp, 1982). Sentences offer the highest level of redundancy due to their syntactic and semantic context, prosodic features, and intonational patterns (e.g., Gardner et al., 1975), which collectively provide multiple cues that can compensate for gaps in spectral resolution (e.g., Shannon et al., 2004). Recognition of isolated words, while still benefiting from lexical and contextual cues, require more precise spectral resolution for accurate recognition, as the listener relies heavily on clear representation of formant transitions and phonemic contrasts within a limited context. Recognition of phonemes, on the other hand, involves the least contextual cues, as they lack contextual or prosodic support and depend almost entirely on distinct spectral and temporal features, such as formant frequencies and their transition over time. This gains particular importance when discussing access to spectral information in CI listeners, which is mainly through access to modulation frequency within and across CI channels. The choice of stimulus can substantially impact test outcomes and its association with spectral resolution ability. Higher redundancy in sentences may underestimate spectral resolution deficits, while phoneme-based tasks can reveal more granular details about individuals’ ability to resolve spectral information. Although testing with sentences better reflects real-life scenarios, more sensitive stimuli such as phonemes are necessary for fine-tuning CI programming, as they provide a more precise assessment of spectral resolution, minimize cognitive compensation, and help optimize electrode stimulation parameters and strategies for a subset of electrodes with poor capacity in transmitting spectral information.

Furthermore, the type of spectral resolution test and the psychophysical and functional aspects it targets can influence its relationship with speech perception. Considering the categorization nature of speech perception (Holt & Lotto, 2010), many traditional spectral resolution assessments rely on non-speech psychophysical tasks that may not reflect this feature (e.g., DeVries & Arenberg, 2018a; Nelson et al., 2011). In contrast, speech-based assessments, such as those using categorization of spectrally manipulated syllables, have been proposed to better capture the functional use of spectral cues in CI listeners (Winn et al., 2015, 2016; Winn & Litovsky, 2015). These tasks minimize semantic and temporal redundancies and assess acoustic-phonetic cue weighting, offering potential sensitivity to subtle perceptual differences that broader speech recognition tasks may overlook. Still, non-speech-based methods remain valuable for isolating basic auditory resolution without the confounding effects of language processing. From this perspective, using tones or modulated signals helps clarify auditory capacity at a more peripheral level. A combination of both may offer the most comprehensive understanding of spectral resolution in CI listeners. Another important methodological consideration, particularly regarding SRD and SMD, is that they may not exclusively measure spectral resolution. Instead, listeners might rely on other cues, such as local loudness differences between the target and reference stimuli (Azadpour & McKay, 2012; Aronoff & Landsberger, 2013; Winn & O’Brien, 2022). This underscores the need to select assessment methods that accurately isolate spectral resolution as a key limitation of CI devices.

It should be noted that performance on many of the spectral resolution measurements and speech perception tasks as well as the results for their link may be affected by individuals’ cortical auditory processing and cognitive abilities (e.g., Farrar et al., 2024; Moberly & Reed, 2019; Mattingly et al., 2018). More specifically, many of the measures discussed involve attention during the task. This is also true in speech in noise perception tasks as results can be significantly impacted by the level of both cognitive processing and efferent inhibitory neural pathways (e.g., Price & Moncrieff, 2021; Beckers et al., 2023; Nemati et al., 2024). Although limited, evidence from studies comparing the speech evoked cortical activities between CI listeners and TH individuals indicate weaker brain activity in bilateral superior temporal gyrus (STG) and middle temporal gyrus (MTG) in temporal lobe (Farrar et al., 2024; Levin et al. 2022; Steinmetzger et al., 2022), which may be attributed to suboptimal spectral resolution in CI listeners (e.g., Arjmandi et al. 2022a; DeVries and Arenberg 2018a; DiNino et al. 2016; Levin et al. 2022; Loizou & Poroy 2001). However, there is very limited evidence on the cortical processing of spectral information and its role on spectral resolution processing on speech perception outcomes (Levin et al., 2022; Steinmetzger et al., 2022; Farrar et al., 2024).

## 5. Conclusions

In conclusion, many spectral resolution measures show promise for predicting speech perception performance in postlingually deafened adult CI listeners. Some approaches, such as SRD, have demonstrated relatively strong associations with speech perception and may serve as reliable tools for assessing global spectral resolution ability. Time-efficient methods such as QSMD and SMRT have further enhanced the clinical feasibility of global measures (Aronoff & Landsberger, 2013; Gifford et al., 2018). The emergence of speech-based tasks as more functional approaches is beneficial for both researchers and clinicians in understanding the connection between spectral resolution and speech perception. However, some weak correlations observed in these studies necessitate further investigation to clarify the underlying mechanisms (e.g., Winn & Litovsky, 2015). The choice of a spectral resolution measure should be guided by its intended application. Global measures are well suited for evaluating the overall effects of speech processing strategies, CI programming, and auditory rehabilitation on spectral resolution and speech recognition. In contrast, channel-specific measures, such as PTCs, although more time-consuming, may be essential for identifying poorly performing electrodes and guiding targeted programming strategies such as electrode deactivation, current steering, or focused stimulation.

For spectral resolution measures to be effective and clinically applicable, they need to satisfy multiple criteria. First, they need to be designed to capture the nature of CI signal processing in coding spectral information in speech and account for modulation frequency within and across spectral bands (Zeng et al., 2008; Loizou, 1998; Dorman & Wolson, 2004). Second, providing channel-specific information about spectral resolution ability might be essential for selectively manipulating CI channels, whether through deactivation, current focusing or current steering (in Advanced Bionic users), to reduce channel interaction, enhance spectral resolution, and ultimately improve speech recognition. Finally, these measures need to be fast enough for clinical application. Some, like the QSMD test (Gifford et al., 2018) and SMRT (Aronoff & Landsberger, 2013), are quick, taking only a few minutes, and are readily available as downloadable software (https://www.ear-lab.org/software-downloads.html). Others, such as PTCs, are channel-specific and labor-intensive, and while earlier studies reported weak or non-significant associations with speech perception (DeVries & Arenberg, 2018a; Anderson et al., 2011; Nelson et al., 2011), more recent evidence suggests that PTCs may capture important aspects of spectral resolution relevant to speech perception under certain listening conditions including in background noise (e.g., Arjmandi et al., 2025).

### 5.1. Future Research Directions

Findings from this review provide valuable insights into future research on assessing spectral resolution and its relationship with speech perception. An important but less frequently addressed topic is about the distinct contributions of peripheral (processing at the level of the cochlea and auditory nerves) and central spectral resolution abilities (processing at higher auditory centers in the brain) to speech perception outcomes. Although distinguishing between the contributions of peripheral and central auditory pathways in the relationship between spectral resolution and speech perception is inherently difficult and methodologically challenging, doing so is crucial for identifying the origins of spectral resolution limitations and understanding their impact on speech understanding. If peripheral spectral resolution is the primary limitation, interventions can focus on improving peripheral limitations such as channel interaction (e.g., DeVries & Arenberg, 2018b; Bierer & Litvak, 2016; Caswell-Midwinter & Arenberg, 2023) and frequency-place mismatch (e.g., Sagi, & Svirsky, 2024). If central spectral resolution plays a larger role, training or rehabilitative therapies targeting neural plasticity could be more effective. Expanding research into the cortical processing of spectral information and speech sounds can provide deeper insights into the neural mechanisms underlying spectral resolution ability and speech perception, individual variability on these abilities, and potential avenues to maximize CI benefits. Lastly, future studies are warranted to examine how individual differences in cognitive abilities may facilitate or hinder the link between spectral resolution and speech perception in CI listeners, providing a better understanding of the factors that explain the connection between spectral resolution ability and speech perception outcomes. In addition, while this review focuses on postlingually deafened adult CI listeners, a future review on children with CIs is warranted. Research on the relationship between spectral resolution and speech recognition in children remains limited (e.g., DeFreese et al., 2024; Arjmandi et al., 2025; Jahn et al., 2022). This is particularly important considering that children with CIs develop largely variable speech recognition abilities (e.g., Arjmandi et al., 2022b; Davidson et al., 2011; Dunn et al., 2014; Tomblin et al., 2005) and language outcomes (Niparko et al., 2010; Svirsky et al., 2004; Arjmandi et al., 2022c; Dilley et al., 2020).

## Data Availability

All data produced in the present study are available upon reasonable request to the authors

## Acknowledgements

We gratefully acknowledge the support provided by startup funds from the University of South Carolina to M.A.; a Presidential Fellowship awarded to S.A.; the Magellan Scholar Grant, awarded to R.F. and K.M., from the Office of the Vice President for Research at USC; and the Honors College Research Grant, awarded to R.F. and K.M. at USC, as well as the assistance of the USC Institute for Mind and Brain in supporting this work.

## Data Availability Statement

Data supporting this manuscript are available upon request from the corresponding author.

## Author Contributions

MA conceptualized the idea and theoretical framework for this review and supervised all aspects of the study. MA and SA contributed to establishing the inclusion/exclusion criteria and the strategy for identifying and selecting literature. MA, SA, RF, MP, LD, and KM conducted the literature search, extracted key information from the included studies, and contributed to the organization and interpretation of the findings. MA and SA wrote the initial version of the manuscript and created the figure and table. All authors contributed to reviewing, revising, and refining the manuscript. MA led the final synthesis and editing of the manuscript. All authors reviewed and approved the final version.

## References

1. Abbas, P. J., Hughes, M. L., Brown, C. J., Miller, C. A., & South, H. (2004). Channel interaction in cochlear implant users evaluated using the electrically evoked compound action potential. Audiology and Neurootology, 9(4), 203–213.

2. Alexander, J. M., & Kluender, K. R. (2008). Spectral tilt change in stop consonant perception. The Journal of the Acoustical Society of America, 123(1), 386–396.

3. Apoux, F., & Healy, E. W. (2012). Use of a compound approach to derive auditory-filter-wide frequency-importance functions for vowels and consonants. The Journal of the Acoustical Society of America, 132(2), 1078–1087.

4. Arenberg Bierer, J. (2010). Probing the electrode-neuron interface with focused cochlear implant stimulation. Trends in amplification, 14(2), 84–95.

5. Arenberg, J. G., Parkinson, W. S., Litvak, L., Chen, C., Kreft, H. A., & Oxenham, A. J. (2018). A dynamically focusing cochlear implant strategy can improve vowel identification in noise. Ear and Hearing, 39(6), 1136–1145.

6. Arenberg J.G., Jahn K.N., Charles H., & Arjmandi M. K. (2022). Psychophysical Tuning Curves as a Measure of Spectral Resolution in Children and Adults with Cochlear Implants. 19th International Symposium on Hearing: Psychoacoustics, Physiology of Hearing, and Auditory Modelling, from the Ear to the Brain (ISH2022), Lyon, France.

7. Arjmandi, M. K., Jahn, K. N., Hem, C. B., & Arenberg, J. G. (2025). Relationship between psychophysical tuning curves and vowel identification in noise in children and adults with cochlear implants. Journal of Speech, Language, and Hearing Research, 1–11.

8. Arjmandi, M. K., Jahn, K. N., & Arenberg, J. G. (2022a). Single-channel focused thresholds relate to vowel identification in pediatric and adult cochlear implant listeners. Trends in Hearing, 26, 23312165221095364.

9. Arjmandi, M., Dilley, L. C., & Wagner, S. E. (2018). Investigation of acoustic dimension use in dialect production: machine learning of sonorant sounds for modeling acoustic cues of African American dialect. In 11th International Conference on Voice Physiology and Biomechanics (Vol. 12, p. 13). USA: East Lansing.

10. Arjmandi, M. K., Herrmann, B. S., Caswell-Midwinter, B., Doney, E. M., & Arenberg, J. G. (2022b). A modified pediatric ranked order speech perception score to assess speech recognition development in children with cochlear implants. American Journal of Audiology, 31(3), 613–632.

11. Arjmandi, M. K., Houston, D., & Dilley, L. C. (2022c). Variability in quantity and quality of early linguistic experience in children with cochlear implants: Evidence from analysis of natural auditory environments. Ear and hearing, 43(2), 685–698.

12. Anderson, E. S., Oxenham, A. J., Nelson, P. B., & Nelson, D. A. (2012). Assessing the role of spectral and intensity cues in spectral ripple detection and discrimination in cochlear- implant users. The Journal of the Acoustical Society of America, 132(6), 3925–3934.

13. Anderson, E. S., Nelson, D. A., Kreft, H., Nelson, P. B., & Oxenham, A. J. (2011). Comparing spatial tuning curves, spectral ripple resolution, and speech perception in cochlear implant users. The Journal of the Acoustical Society of America, 130(1), 364–375.

14. Aronoff, J. M., & Landsberger, D. M. (2013). The development of a modified spectral ripple test. The Journal of the Acoustical Society of America, 134(2), EL217–EL222.

15. Ashjaei, S., Jalilvand, H., Hadipour, H., & Sadeghi, R. (2022). Acceptable Noise Level in Children with Unilateral Cochlear Implant and Bimodal Hearing. Auditory and Vestibular Research.

16. Ashjaei, S., Behroozmand, R., Fozdar, S., Farrar, R., & Arjmandi, M. (2024). Vocal Control and Speech Production in Cochlear Implant Listeners: A Review within Auditory-Motor Processing Framework. Hearing Research, 109132.

17. Azadpour, M., & McKay, C. M. (2012). A psychophysical method for measuring spatial resolution in cochlear implants. Journal of the Association for Research in Otolaryngology, 13, 145–157.

18. Azadpour, M., McKay, C. M., & Svirsky, M. A. (2018). Effect of pulse rate on loudness discrimination in cochlear implant users. Journal of the Association for Research in Otolaryngology, 19, 287–299.

19. Baer, T., & Moore, B. C. (1993). Effects of spectral smearing on the intelligibility of sentences in noise. The Journal of the Acoustical Society of America, 94(3), 1229–1241.

20. Bálint, A., Wimmer, W., Rummel, C., Caversaccio, M., & Weder, S. (2024, July). Neural Correlates of Speech Comprehension in Normal Hearing Individuals and Cochlear Implant Users-An fNIRS Study in Quiet and Noisy Environments. In 2024 46th Annual International Conference of the IEEE Engineering in Medicine and Biology Society (EMBC) (pp. 1–5). IEEE.

21. Bálint, A., Wimmer, W., Rummel, C., Caversaccio, M., & Weder, S. (2024). Neural Correlates of Speech Comprehension in Normal Hearing Individuals and Cochlear Implant Users-An fNIRS Study in Quiet and Noisy Environments. In 2024 46th Annual International Conference of the IEEE Engineering in Medicine and Biology Society (EMBC) (pp. 1–5). IEEE.

22. Berenstein, C. K., Mens, L. H., Mulder, J. J., & Vanpoucke, F. J. (2008). Current steering and current focusing in cochlear implants: Comparison of monopolar, tripolar, and virtual channel electrode configurations. Ear and Hearing, 29(2), 250–260.

23. Başkent, D., and Shannon, R. V. (2006). “Frequency transposition around dead regions simulated with a noise band vocoder,” Journal of Acoustic Society of America. 119(2), 1156–1163.

24. Beckers, L., Tromp, N., Philips, B., Mylanus, E., & Huinck, W. (2023). Exploring neurocognitive factors and brain activation in adult cochlear implant recipients associated with speech perception outcomes—A scoping review. Frontiers in Neuroscience, 17, 1046669.

25. Bierer, J. A., & Litvak, L. (2016). Reducing channel interaction through cochlear implant programming may improve speech perception: Current focusing and channel deactivation. Trends in Hearing, 20, 233121651665338.

26. Boëx, C., de Balthasar, C., Kós, M. I., & Pelizzone, M. (2003). Electrical field interactions in different cochlear implant systems. The Journal of the Acoustical Society of America, 114(4), 2049–2057.

27. Busby, P. A., Tong, Y. C., & Clark, G. M. (1993). Electrode position, repetition rate, and speech perception by early- and late-deafened cochlear implant patients. Journal of the Acoustical Society of America, 93(2), 1058–1067.

28. Canfarotta, M. W., Dillon, M. T., Buss, E., Pillsbury, H. C., Brown, K. D., & O’Connell, B. P. (2020). Frequency-to-place mismatch: characterizing variability and the influence on speech perception outcomes in cochlear implant recipients. Ear and hearing, 41(5), 1349–1361.

29. Caswell-Midwinter, B., Doney, E. M., Arjmandi, M. K., Jahn, K. N., Herrmann, B. S., & Arenberg, J. G. (2022). The relationship between impedance, programming and word recognition in a large clinical dataset of cochlear implant recipients. Trends in Hearing, 26, 23312165211060983.

30. Caswell-Midwinter, B., & Arenberg, J. G. (2023). Comparing Fixed and Individualized Channel Interaction Coefficients for Speech Perception With Dynamic Focusing Cochlear Implant Strategies. Trends in Hearing, 27, 23312165231176157.

31. Christensen, L. A., & Humes, L. E. (1997). Identification of multidimensional stimuli containing speech cues and the effects of training. The Journal of the Acoustical Society of America, 102(4), 2297–2310.

32. Cucis, P. A., Berger-Vachon, C., Thaï-Van, H., Hermann, R., Gallego, S., & Truy, E. (2021). Word recognition and frequency selectivity in cochlear implant simulation: Effect of channel interaction. Journal of Clinical Medicine, 10(4), 679.

33. Cychosz, M., Winn, M. B., & Goupell, M. J. (2024). How to vocode: Using channel vocoders for cochlear-implant research. The Journal of the Acoustical Society of America, 155(4), 2407–2437.

34. Davidson, L. S., Geers, A. E., Blamey, P. J., Tobey, E., & Brenner, C. (2011). Factors contributing to speech perception scores in long-term pediatric cochlear implant users. Ear and Hearing, 32(1), 19S–26S.

35. DeVries, L., & Arenberg, J. G. (2018a). Psychophysical tuning curves as a correlate of electrode position in cochlear implant listeners. Journal of the Association for Research in Otolaryngology, 19, 571–587.

36. DeVries, L., & Arenberg, J. G. (2018b). Current focusing to reduce channel interaction for distant electrodes in cochlear implant programs. Trends in hearing, 22, 2331216518813811.

37. DeFreese, A., Camarata, S., Sunderhaus, L., Holder, J., Berg, K., Lighterink, M., & Gifford, R. (2024). The impact of spectral and temporal processing on speech recognition in children with cochlear implants. Scientific Reports, 14(1), 14094.

38. de Jong, M. A., Briaire, J. J., & Frijns, J. H. (2019). Dynamic current focusing: a novel approach to loudness coding in cochlear implants. Ear and Hearing, 40(1), 34–44.

39. Dilley, L., Lehet, M., Wieland, E. A., Arjmandi, M. K., Kondaurova, M., Wang, Y., … & Bergeson, T. (2020). Individual differences in mothers’ spontaneous infant-directed speech predict language attainment in children with cochlear implants. Journal of Speech, Language, and Hearing Research, 63(7), 2453–2467.

40. DiNino, M., O’Brien, G., Bierer, S. M., Jahn, K. N., & Arenberg, J. G. (2019). The estimated electrode-neuron interface in cochlear implant listeners is different for early-implanted children and late-implanted adults. Journal of the Association for Research in Otolaryngology, 20, 291–303.

41. DiNino, M., Wright, R. A., Winn, M. B., & Bierer, J. A. (2016). Vowel and consonant confusions from spectrally manipulated stimuli designed to simulate poor cochlear implant electrode- neuron interfaces. The Journal of the Acoustical Society of America, 140(6), 4404–4418.

42. DiNino, M., Arenberg, J. G., Duchen, A. L., & Winn, M. B. (2020). Effects of age and cochlear implantation on spectrally cued speech categorization. Journal of Speech, Language, and Hearing Research, 63(7), 2425–2440.

43. Donaldson, G. S., Rogers, C. L., Cardenas, E. S., Russell, B. A., & Hanna, N. H. (2013). Vowel identification by cochlear implant users: Contributions of static and dynamic spectral cues. The Journal of the Acoustical Society of America, 134(4), 3021–3028.

44. Donaldson, G. S., Rogers, C. L., Johnson, L. B., & Oh, S. H. (2015). Vowel identification by cochlear implant users: Contributions of duration cues and dynamic spectral cues. The Journal of the Acoustical Society of America, 138(1), 65–73.

45. Donaldson, G. S., & Nelson, D. A. (2000). Place-pitch sensitivity and its relation to consonant recognition by cochlear implant listeners using the MPEAK and SPEAK speech processing strategies. The Journal of the Acoustical Society of America, 107(3), 1645–1658.

46. Dorman, M. F., & Wilson, B. S. (2004). The design and function of cochlear implants: fusing medicine, neural science and engineering, these devices transform human speech into an electrical code that deafened ears can understand. American Scientist, 92(5), 436–445.

47. Dorman, M. F., & Loizou, P. C. (1996). Relative spectral change and formant transitions as cues to labial and alveolar place of articulation. The Journal of the Acoustical Society of America, 100(6), 3825–3830.

48. Drennan, W. R., Anderson, E. S., Won, J. H., & Rubinstein, J. T. (2014). Validation of a clinical assessment of spectral-ripple resolution for cochlear implant users. Ear and hearing, 35(3), e92–e98.

49. Dunn, C. C., Walker, E. A., Oleson, J., Kenworthy, M., Van Voorst, T., Tomblin, J. B., Ji, H., Kirk, K., McMurray, B., Hanson, M., & Gantz, B. J. (2014). Longitudinal speech perception and language performance in pediatric cochlear implant users: The effect of age at implantation. Ear and Hearing, 35(2), 148–160

50. Farrar, R., Ashjaei, S., & Arjmandi, M. K. (2024). Speech-evoked cortical activities and speech recognition in adult cochlear implant listeners: a review of functional near-infrared spectroscopy studies. Experimental Brain Research, 1-22.

51. Feng, L., & Oxenham, A. J. (2018). Effects of spectral resolution on spectral contrast effects in cochlear-implant users. The Journal of the Acoustical Society of America, 143(6), EL468–EL473.

52. Finke, M., Büchner, A., Ruigendijk, E., Meyer, M., & Sandmann, P. (2016). On the relationship between auditory cognition and speech intelligibility in cochlear implant users: An ERP study. Neuropsychologia, 87, 169–181.

53. Fleming, J. T., & Winn, M. B. (2022). Strategic perceptual weighting of acoustic cues for word stress in listeners with cochlear implants, acoustic hearing, or simulated bimodal hearing. The Journal of the Acoustical Society of America, 152(3), 1300–1316.

54. Friesen, L. M., Shannon, R. V., Baskent, D., & Wang, X. (2001). Speech recognition in noise as a function of the number of spectral channels: Comparison of acoustic hearing and cochlear implants. The Journal of the Acoustical Society of America, 110(2), 1150–1163.

55. Fu, Q. J., & Nogaki, G. (2005). Noise susceptibility of cochlear implant users: the role of spectral resolution and smearing. Journal of the Association for Research in Otolaryngology, 6, 19–27.

56. Gardner, H., Albert, M. L., & Weintraub, S. (1975). Comprehending a word: The influence of speed and redundancy on auditory comprehension in aphasia. Cortex, 11(2), 155–162.

57. Geurts, L., & Wouters, J. (2004). Better place-coding of the fundamental frequency in cochlear implants. The Journal of the Acoustical Society of America, 115(2), 844–852.

58. Gifford, R. H., Noble, J. H., Camarata, S. M., Sunderhaus, L. W., Dwyer, R. T., Dawant, B. M., … & Labadie, R. F. (2018). The relationship between spectral modulation detection and speech recognition: Adult versus pediatric cochlear implant recipients. Trends in hearing, 22, 2331216518771176.

59. Gifford, R. H., Hedley-Williams, A., & Spahr, A. J. (2014). Clinical assessment of spectral modulation detection for adult cochlear implant recipients: A non-language based measure of performance outcomes. International journal of audiology, 53(3), 159–164.

60. Goehring, T., Archer-Boyd, A. W., Arenberg, J. G., & Carlyon, R. P. (2021). The effect of increased channel interaction on speech perception with cochlear implants. Scientific Reports, 11(1), 10383.

61. Goehring, T., Arenberg, J. G., & Carlyon, R. P. (2020). Using spectral blurring to assess effects of channel interaction on speech-in-noise perception with cochlear implants. Journal of the Association for Research in Otolaryngology, 21(4), 353–371.

62. Goldwyn, J. H., Bierer, S. M., & Bierer, J. A. (2010). Modeling the electrode–neuron interface of cochlear implants: Effects of neural survival, electrode placement, and the partial tripolar configuration. Hearing research, 268(1-2), 93–104.

63. Gonzalez, J., & Oliver, J. C. (2005). Gender and speaker identification as a function of the number of channels in spectrally reduced speech. The Journal of the Acoustical Society of America, 118(1), 461–470.

64. Goupell, M. J. (2015). Pushing the envelope of auditory research with cochlear implants. Acoustics Today, 11(2), 26–33.

65. Goycoolea, M. V., Muchow, D. C., Schirber, C. M., Goycoolea, H. G., & Schellhas, K. (1990). Anatomical perspective, approach, and experience with multichannel intracochlear implantation. The Laryngoscope, 100(S50), 1–18.

66. Green, K. M., Bhatt, Y. M., Mawman, D. J., O’Driscoll, M. P., Saeed, S. R., Ramsden, R. T., & Green, M. W. (2007). Predictors of audiological outcome following cochlear implantation in adults. Cochlear implants international, 8(1), 1–11.

67. Harmon, T. G., Dromey, C., Nelson, B., & Chapman, K. (2021). Effects of Background Noise on Speech and Language in Young Adults. Journal of speech, language, and hearing research, 64(4), 1104–1116.

68. Hedrick, M. S., & Carney, A. E. (1997). Effect of relative amplitude and formant transitions on perception of place of articulation by adult listeners with cochlear implants. Journal of Speech, Language, and Hearing Research, 40(6), 1445–1457.

69. Henry, B. A., & Turner, C. W. (2003). The resolution of complex spectral patterns by cochlear implant and normal-hearing listeners. The Journal of the Acoustical Society of America, 113(5), 2861–2873.

70. Henry, B. A., Turner, C. W., & Behrens, A. (2005). Spectral peak resolution and speech recognition in quiet: Normal hearing, hearing impaired, and cochlear implant listeners. The Journal of the Acoustical Society of America, 118(2), 1111–1121.

71. Hillenbrand, J., Getty, L. A., Clark, M. J., & Wheeler, K. (1995). Acoustic characteristics of American English vowels. The Journal of the Acoustical Society of America, 97(5), 3099– 3111.

72. Holden, L. K., Finley, C. C., Firszt, J. B., Holden, T. A., Brenner, C., Potts, L. G., … & Skinner, M. W. (2013). Factors affecting open-set word recognition in adults with cochlear implants. Ear and hearing, 34(3), 342–360.

73. Holden, L. K., Firszt, J. B., Reeder, R. M., Uchanski, R. M., Dwyer, N. Y., & Holden, T. A. (2016). Factors affecting outcomes in cochlear implant recipients implanted with a perimodiolar electrode array located in scala tympani. Otology & Neurotology, 37(10), 1662–1668.

74. Holmes, A. E., Kemker, E. J., & Merwin, G. E. (1987). The effects of varying the number of cochlear implant electrodes on speech perception. Otology & Neurotology, 8(3), 240–246.

75. Holt, L. L., & Lotto, A. J. (2006). Cue weighting in auditory categorization: Implications for first and second language acquisition. The Journal of the Acoustical Society of America, 119(5), 3059–3071.

76. Holt, L. L., & Lotto, A. J. (2010). Speech perception as categorization. *Attention, Perception*, & Psychophysics, 72(5), 1218–1227.

77. Horn, D. L., Dudley, D. J., Dedhia, K., Nie, K., Drennan, W. R., Won, J. H., … & Werner, L. A. (2017). Effects of age and hearing mechanism on spectral resolution in normal hearing and cochlear-implanted listeners. The Journal of the Acoustical Society of America, 141(1), 613–623.

78. Hughes, M. L., & Stille, L. J. (2008). Psychophysical versus physiological spatial forward masking and the relation to speech perception in cochlear implants. Ear and hearing, 29(3), 435–452.

79. Jahn, K. N., & Arenberg, J. G. (2020). Electrophysiological estimates of the electrode–neuron interface differ between younger and older listeners with cochlear implants. Ear and Hearing, 41(4), 948–960.

80. Jahn, K. N., Arenberg, J. G., & Horn, D. L. (2022). Spectral resolution development in children with normal hearing and with cochlear implants: A review of behavioral studies. *Journal of Speech*, Language, and Hearing Research, 65(4), 1646–1658.

81. Jahn, K. N., & Arenberg, J. G. (2020). Identifying cochlear implant channels with relatively poor electrode-neuron interfaces using the electrically evoked compound action potential. Ear and Hearing, 41(4), 961–973.

82. Jeon, E. K., Turner, C. W., Karsten, S. A., Henry, B. A., & Gantz, B. J. (2015). Cochlear implant users’ spectral ripple resolution. The Journal of the Acoustical Society of America, 138(4), 2350–2358.

83. Jones, G. L., Ho Won, J., Drennan, W. R., & Rubinstein, J. T. (2013). Relationship between channel interaction and spectral-ripple discrimination in cochlear implant users. The Journal of the Acoustical Society of America, 133(1), 425–433.

84. Kamakura, T., & Nadol Jr, J. B. (2016). Correlation between word recognition score and intracochlear new bone and fibrous tissue after cochlear implantation in the human. Hearing research, 339, 132–141.

85. Kwon, B. J., & van den Honert, C. (2006). Effect of electrode configuration on psychophysical forward masking in cochlear implant listeners. The Journal of the Acoustical Society of America, 119(5), 2994–3002.

86. Landsberger, D. M., Padilla, M., Martinez, A. S., & Eisenberg, L. S. (2018). Spectral-temporal modulated ripple discrimination by children with cochlear implants. Ear and Hearing, 39(1), 60–68.

87. Landsberger, D. M., Stupak, N., & Aronoff, J. M. (2019). Spectral-temporally modulated ripple test lite for computeRless measurement (SLRM): A nonlinguistic test for audiology clinics. Ear and hearing, 40(5), 1253–1255.

88. Lawler, M., Yu, J., & Aronoff, J. M. (2017). Comparison of the spectral-temporally modulated ripple test with the Arizona Biomedical Institute Sentence Test in cochlear implant users. Ear and hearing, 38(6), 760–766.

89. Lehiste, I. (1965). Acoustical Characteristics of Selected English Consonants. Language, 41, 332.

90. Leake, P. A., & Hradek, G. T. (1988). Cochlear pathology of long term neomycin induced deafness in cats. Hearing research, 33(1), 11–33.

91. Levin, M., Balberg, M., & Zaltz, Y. (2022). Cortical activation in response to speech differs between prelingually deafened cochlear implant users with good or poor speech-in-noise understanding: an fNIRS study. Applied Sciences, 12(23), 12063.

92. Lin, Y. S., Lu, H. P., Hung, S. C., & Chang, C. P. (2009). Lexical tone identification and consonant recognition in acoustic simulations of cochlear implants. Acta oto-laryngologica, 129(6), 630–637.

93. Liepins, R., Kaider, A., Honeder, C., Auinger, A. B., Dahm, V., Riss, D., & Arnoldner, C. (2020). Formant frequency discrimination with a fine structure sound coding strategy for cochlear implants. Hearing Research, 392, 107970.

94. Litvak, L. M., Spahr, A. J., Saoji, A. A., & Fridman, G. Y. (2007). Relationship between perception of spectral ripple and speech recognition in cochlear implant and vocoder listeners. The Journal of the Acoustical Society of America, 122(2), 982–991.

95. Loizou, P. C. (1998). Mimicking the human ear. IEEE signal processing magazine, 15(5), 101–130.

96. Loizou, P. C., Dorman, M., & Tu, Z. (1999). On the number of channels needed to understand speech. The Journal of the Acoustical Society of America, 106(4), 2097–2103.

97. Loizou, P. C., & Poroy, O. (2001). Minimum spectral contrast needed for vowel identification by normal hearing and cochlear implant listeners. The Journal of the Acoustical Society of America, 110(3), 1619–1627.

98. Long, C. J., Holden, T. A., McClelland, G. H., Parkinson, W. S., Shelton, C., Kelsall, D. C., & Smith, Z. M. (2014). Examining the electro-neural interface of cochlear implant users using psychophysics, CT scans, and speech understanding. Journal of the Association for Research in Otolaryngology, 15, 293–304.

99. Lorenzi, C., Gilbert, G., Carn, H., Garnier, S., & Moore, B. C. (2006). Speech perception problems of the hearing impaired reflect inability to use temporal fine structure. Proceedings of the National Academy of Sciences, 103(49), 18866–18869.

100. Mattingly, J. K., Castellanos, I., & Moberly, A. C. (2018). Nonverbal reasoning as a contributor to sentence recognition outcomes in adults with cochlear implants. Otology & Neurotology, 39(10), e956–e963.

101. Mehta, A. H., Lu, H., & Oxenham, A. J. (2020). The Perception of Multiple Simultaneous Pitches as a Function of Number of Spectral Channels and Spectral Spread in a Noise- Excited Envelope Vocoder. Journal of the Association for Research in Otolaryngology, 21(1), 61–72.

102. Mehta, A. H., & Oxenham, A. J. (2017). Vocoder Simulations Explain Complex Pitch Perception Limitations Experienced by Cochlear Implant Users. Journal of the Association for Research in Otolaryngology, 18(6), 789–802.

103. McArdle, R., & Wilson, R. H. (2009). Speech perception in noise: The basics. Perspectives on hearing and hearing disorders: research and diagnostics, 13(1), 4–13.

104. Moberly, A. C., Lowenstein, J. H., Tarr, E., Caldwell-Tarr, A., Welling, D. B., Shahin, A. J., & Nittrouer, S. (2014). Do adults with cochlear implants rely on different acoustic cues for phoneme perception than adults with normal hearing?. Journal of Speech, Language, and Hearing Research, 57(2), 566–582.

105. Moberly, A. C., & Reed, J. (2019). Making sense of sentences: Top-down processing of speech by adult cochlear implant users. Journal of Speech, Language, and Hearing Research, 62(8), 2895–2905.

106. Moberly, A. C., Vasil, K., Baxter, J., Klamer, B., Kline, D., & Ray, C. (2020). Comprehensive auditory rehabilitation in adults receiving cochlear implants: A pilot study. Laryngoscope investigative otolaryngology, 5(5), 911–918.

107. Moore, B. C. (1978). Psychophysical tuning curves measured in simultaneous and forward masking. The Journal of the Acoustical Society of America, 63(2), 524–532.

108. Nelson, D. A., & Freyman, R. L. (1984). Broadened forward masked tuning curves from intense masking tones: Delay time and probe level manipulations. The Journal of the Acoustical Society of America, 75(5), 1570–1577.

109. Nelson, D. A., Van Tasell, D. J., Schroder, A. C., Soli, S., & Levine, S. (1995). Electrode ranking of ‘‘place pitch’’and speech recognition in electrical hearing. The Journal of the Acoustical Society of America, 98(4), 1987–1999.

110. Nelson, D. A., Donaldson, G. S., & Kreft, H. (2008). Forward-masked spatial tuning curves in cochlear implant users. The Journal of the Acoustical Society of America, 123(3), 1522–1543.

111. Nelson, D. A., Kreft, H. A., Anderson, E. S., & Donaldson, G. S. (2011). Spatial tuning curves from apical, middle, and basal electrodes in cochlear implant users. The Journal of the Acoustical Society of America, 129(6), 3916–3933.

112. Nie, K., Barco, A., & Zeng, F. G. (2006). Spectral and temporal cues in cochlear implant speech perception. Ear and hearing, 27(2), 208–217.

113. Nie, K., Stickney, G., & Zeng, F. G. (2005). Encoding frequency modulation to improve cochlear implant performance in noise. IEEE Transactions on Biomedical Engineering, 52(1), 64–73.

114. Niparko, J. K., Tobey, E. A., Thal, D. J., Eisenberg, L. S., Wang, N. Y., Quittner, A. L., … & CDaCI Investigative Team. (2010). Spoken language development in children following cochlear implantation. Jama, 303(15), 1498–1506.

115. Noble, J. H., Gifford, R. H., Hedley-Williams, A. J., Dawant, B. M., & Labadie, R. F. (2014). Clinical evaluation of an image-guided cochlear implant programming strategy. Audiology and Neurotology, 19(6), 400–411.

116. Noble, J. H., Hedley-Williams, A. J., Sunderhaus, L., Dawant, B. M., Labadie, R. F., Camarata, S. M., & Gifford, R. H. (2016). Initial results with image-guided cochlear implant programming in children. Otology & Neurotology, 37(2), e63–e69.

117. Noble, J. H., Labadie, R. F., Gifford, R. H., & Dawant, B. M. (2013). Image-guidance enables new methods for customizing cochlear implant stimulation strategies. IEEE Transactions on Neural Systems and Rehabilitation Engineering, 21(5), 820–829

118. O’Donoghue, G. (2013). Cochlear implants—science, serendipity, and success. N Engl J Med, 369(13), 1190–1193.

119. O’Neill, E. R., Kreft, H. A., & Oxenham, A. J. (2019). Speech perception with spectrally non- overlapping maskers as measure of spectral resolution in cochlear implant users. Journal of the Association for Research in Otolaryngology, 20(2), 151–167.

120. Otte J., Schuknecht H. F., & Kerr A. G. (1978). Ganglion cell populations in normal and pathological human, implications for cochlear implantation. The Laryngoscope, 88(8), 1231–1246.

121. Oxenham, A. J., & Kreft, H. A. (2014). Speech perception in tones and noise via cochlear implants reveals influence of spectral resolution on temporal processing. Trends in Hearing, 18, 2331216514553783.

122. Peterson, G. E., & Lehiste, I. (1962). Revised CNC lists for auditory tests. The Journal of speech and hearing disorders, 27, 62–70.

123. Peng, S. C., Chatterjee, M., & Lu, N. (2012). Acoustic cue integration in speech intonation recognition with cochlear implants. Trends in Amplification, 16(2), 67–82.

124. Price, C. N., & Moncrieff, D. (2021). Defining the Role of Attention in Hierarchical Auditory Processing. Audiology research, 11(1), 112–128.

125. Repp, B. H. (1982). Phonetic trading relations and context effects: New experimental evidence for a speech mode of perception. Psychological bulletin, 92(1), 81.

126. Repp, B. H. (1984). The role of release bursts in the perception of [s]-stop clusters. The Journal of the Acoustical Society of America, 75(4), 1219–1230.

127. Rødvik, A. K., von Koss Torkildsen, J., Wie, O. B., Storaker, M. A., & Silvola, J. T. (2018). Consonant and vowel identification in cochlear implant users measured by nonsense words: A systematic review and meta-analysis. Journal of Speech, Language, and Hearing Research, 61(4), 1023–1050.

128. Rosenblum, L. D. (2008). Speech perception as a multimodal phenomenon. Current directions in psychological science, 17(6), 405–409.

129. Sagi, E., & Svirsky, M. A. (2017). Contribution of formant frequency information to vowel perception in steady-state noise by cochlear implant users. The Journal of the Acoustical Society of America, 141(2), 1027–1038.

130. Sagi, E., & Svirsky, M. A. (2024). A Level-Adjusted Cochlear Frequency-to-Place Map for Estimating Tonotopic Frequency Mismatch With a Cochlear Implant. Ear and Hearing, 10–1097.

131. Saoji, A. A., Litvak, L., Spahr, A. J., & Eddins, D. A. (2009). Spectral modulation detection and vowel and consonant identifications in cochlear implant listeners. The Journal of the Acoustical Society of America, 126(3), 955–958.

132. Scheperle, R. A., & Abbas, P. J. (2015). Peripheral and central contributions to cortical responses in cochlear implant users. Ear and hearing, 36(4), 430–440.

133. Scheperle, R. A., & Abbas, P. J. (2015). Relationships among peripheral and central electrophysiological measures of spatial and spectral selectivity and speech perception in cochlear implant users. Ear and Hearing, 36(4), 441–453.

134. Shannon, R. V. (1990). Forward masking in patients with cochlear implants. The Journal of the Acoustical Society of America, 88(2), 741–744.

135. Shannon, R. V., Zeng, F. G., Kamath, V., Wygonski, J., & Ekelid, M. (1995). Speech recognition with primarily temporal cues. Science, 270(5234), 303–304.

136. Shannon, R. V., Galvin III, J. J., & Baskent, D. (2001). Holes in hearing. Journal of the Association for Research in Otolaryngology, 3(2), 185.

137. Shannon, R. V., Fu, Q. J., Galvin, J., & Friesen, L. (2004). Speech perception with cochlear implants. In Cochlear implants: Auditory prostheses and electric hearing (pp. 334–376). New York, NY: Springer New York.

138. Skinner, M. W., Clark, G. M., Whitford, L. A., Seligman, P. M., Staller, S. J., Shipp, D. B., … & Beiter, A. L. (1994). Evaluation of a new spectral peak coding strategy for the Nucleus 22 channel cochlear implant system. Otology & Neurotology, 15, 15–27.

139. Skinner, M. W., Holden, L. K., Whitford, L. A., Plant, K. L., Psarros, C., & Holden, T. A. (2002). Speech recognition with the nucleus 24 SPEAK, ACE, and CIS speech coding strategies in newly implanted adults. Ear and hearing, 23(3), 207–223.

140. Singh, N. C., & Theunissen, F. E. (2003). Modulation spectra of natural sounds and ethological theories of auditory processing. The Journal of the Acoustical Society of America, 114(6), 3394–3411.

141. Srinivasan, A. G., Padilla, M., Shannon, R. V., & Landsberger, D. M. (2013). Improving speech perception in noise with current focusing in cochlear implant users. Hearing Research, 299, 29–36.

142. Steinmetzger, K., Meinhardt, B., Praetorius, M., Andermann, M., & Rupp, A. (2022). A direct comparison of voice pitch processing in acoustic and electric hearing. NeuroImage: Clinical, 36, 103188.

143. Supin, A. Y., Popov, V. V., Milekhina, O. N., & Tarakanov, M. B. (1994). Frequency resolving power measured by rippled noise. Hearing Research, 78(1), 31–40.

144. Supin, A. Y., Popov, V. V., Milekhina, O. N., & Tarakanov, M. B. (1997). Frequency-temporal resolution of hearing measured by rippled noise. Hearing Research, 108(1-2), 17–27.

145. Supin, A. Y., Popov, V. V., Milekhina, O. N., & Tarakanov, M. B. (1999). Ripple depth and density resolution of rippled noise. The Journal of the Acoustical Society of America, 106(5), 2800–2804.

146. Svirsky, M. A., Robbins, A. M., Kirk, K. I., Pisoni, D. B., & Miyamoto, R. T. (2000). Language development in profoundly deaf children with cochlear implants. Psychological science, 11(2), 153–158.

147. Svirsky, M. (2017). Cochlear implants and electronic hearing. Physics Today, 70(8), 52–58.

148. Svirsky, M. A., Sagi, E., Meyer, T. A., Kaiser, A. R., & Teoh, S. W. (2011). A mathematical model of medial consonant identification by cochlear implant users. The Journal of the Acoustical Society of America, 129(4), 2191–2200.

149. Syrdal, A. K. (1983). Perception of consonant place of articulation. Speech and Language, 9, 313–349.

150. Tomblin, J. B., Barker, B. A., Spencer, L. J., Zhang, X., & Gantz, B. J. (2005). The effect of age at cochlear implant initial stimulation on expressive language growth in infants and toddlers. *Journal of Speech*, Language, and Hearing Research, 48(4), 853–867.

151. Teoh, S. W., Neuburger, H. S., & Svirsky, M. A. (2003). Acoustic and electrical pattern analysis of consonant perceptual cues used by cochlear implant users. Audiology and Neurotology, 8(5), 269–285.

152. Ter Keurs, M., Festen, J. M., & Plomp, R. (1993). Effect of spectral envelope smearing on speech reception. II. The Journal of the Acoustical Society of America, 93(3), 1547–1552.

153. Throckmorton, C. S., & Collins, L. M. (2002). The effect of channel interactions on speech recognition in cochlear implant subjects: Predictions from an acoustic model. The Journal of the Acoustical Society of America, 112(1), 285–296.

154. Townshend, B., Cotter, N., Van Compernolle, D., & White, R. L. (1987). Pitch perception by cochlear implant subjects. The Journal of the Acoustical Society of America, 82(1), 106–115.

155. Toscano, J. C., & McMurray, B. (2010). Cue integration with categories: Weighting acoustic cues in speech using unsupervised learning and distributional statistics. Cognitive science, 34(3), 434–464.

156. Wanna, G. B., Noble, J. H., Carlson, M. L., Gifford, R. H., Dietrich, M. S., Haynes, D. S., … & Labadie, R. F. (2014). Impact of electrode design and surgical approach on scalar location and cochlear implant outcomes. The Laryngoscope, 124(S6), S1–S7.

157. Wilk, M., Hessler, R., Mugridge, K., Jolly, C., Fehr, M., Lenarz, T., & Scheper, V. (2016). Impedance changes and fibrous tissue growth after cochlear implantation are correlated and can be reduced using a dexamethasone eluting electrode. PloS one, 11(2), e0147552.

158. Winn, M. B., & Litovsky, R. Y. (2015). Using speech sounds to test functional spectral resolution in listeners with cochlear implants. The Journal of the Acoustical Society of America, 137(3), 1430–1442.

159. Winn, M. B., Won, J. H., & Moon, I. J. (2016). Assessment of spectral and temporal resolution in cochlear implant users using psychoacoustic discrimination and speech cue categorization. Ear and hearing, 37(6), e377–e390.

160. Winn, M.B. (2020). Accommodation of gender-related phonetic differences by listeners with cochlear implants and in a variety of vocoder simulations. The Journal of the Acoustical Society of America, 147 1, 174 .

161. Winn, M. B., & O’Brien, G. (2022). Distortion of spectral ripples through cochlear implants has major implications for interpreting performance scores. Ear and hearing, 43(3), 764–772.

162. Winn, M. B., Chatterjee, M., & Idsardi, W. J. (2012). The use of acoustic cues for phonetic identification: effects of spectral degradation and electric hearing. The Journal of the Acoustical Society of America, 131(2), 1465–1479.

163. White, M. W., Merzenich, M. M., & Gardi, J. N. (1984). Multichannel cochlear implants: Channel interactions and processor design. Archives of Otolaryngology, 110(8), 493–501.

164. Won, J. H., Drennan, W. R., & Rubinstein, J. T. (2007). Spectral-ripple resolution correlates with speech reception in noise in cochlear implant users. Journal of the Association for Research in Otolaryngology, 8(3), 384–392.

165. Won, J. H., Moon, I. J., Jin, S., Park, H., Woo, J., Cho, Y. S., … & Hong, S. H. (2015). Spectrotemporal modulation detection and speech perception by cochlear implant users. PloS one, 10(10), e0140920.

166. Wouters, J., McDermott, H. J., & Francart, T. (2015). Sound coding in cochlear implants: From electric pulses to hearing. IEEE Signal Processing Magazine, 32(2), 67–80.

167. Xu, K., Willis, S., Gopen, Q., & Fu, Q. J. (2020). Effects of spectral resolution and frequency mismatch on speech understanding and spatial release from masking in simulated bilateral cochlear implants. Ear and hearing, 41(5), 1362–1371.

168. Xu, L., Thompson, C. S., & Pfingst, B. E. (2005). Relative contributions of spectral and temporal cues for phoneme recognition. The Journal of the Acoustical Society of America, 117(5), 3255–3267.

169. Zeng, F. G. (2004). Trends in cochlear implants. Trends in amplification, 8(1), 1–34.

170. Zeng, F. G., Rebscher, S., Harrison, W., Sun, X., & Feng, H. (2008). Cochlear Implants: System Design, Integration, and Evaluation. IEEE Reviews in Biomedical Engineering, 1, 115–142.

171. Zhou, N., Xu, L., & Lee, C. Y. (2010). The effects of frequency-place shift on consonant confusion in cochlear implant simulations. The Journal of the Acoustical Society of America, 128(1), 401–409.

172. Zhu, Z., Tang, Q., Zeng, F. G., Guan, T., & Ye, D. (2012). Cochlear-implant spatial selectivity with monopolar, bipolar and tripolar stimulation. Hearing research, 283(1-2), 45–58.

173. Zwolan, T. A., Collins, L. M., & Wakefield, G. H. (1997). Electrode discrimination and speech recognition in postlingually deafened adult cochlear implant subjects. The Journal of the Acoustical Society of America, 102(6), 3673–3685.

