## Supplementary material for "From Spectral Resolution to Speech Perception: A Review of Findings in Postlingually Deafened Adult Cochlear Implant Listeners": Table 1

**Table 1.** Summary of the findings from prior studies investigating the relationship between measures of spectral resolution and speech recognition in postlingually deafened adult CI listeners. CI: cochlear implant, TH: typical hearing. yo: years old, *R^2^*: coefficient of determination, SNR: signal-to-noise ratio.

| **Study** | **Population** | **Spectral resolution measurement(s)** | **Speech perception measurement**  **(s)** | **Main results**  **(*R^2^*)** |
| --- | --- | --- | --- | --- |
| Arjmandi et al., 2025 | CI listeners (n = 8) (Mean age at CI = 54.25 yo) (Mean duration of CI experience = 7.99 years) | Psychophysical tuning curve (PTCs) | Vowel identification using /hVd/ contexts in multi-talker babble noise with +10 dB SNR | Significant correlation between PTC and vowel identification (0.21). |
| Dinino et al., 2020 | CI listeners (n = 15) and TH (N = 17) (Mean age at CI = 59 yo) (Mean duration of CI experience = 9 years) | Consonant categorization using /ba/-/da/ continuum, /ra/-/la/ and /sa/-/ʃa/ continuum. | Vowel identification using /hVd/ contexts in quiet | Significant correlation between the use of formant transition cue and consonant categorization (No R^2^ reported) |
|  |  | Spectral-Temporally Modulated Ripple Test (SMRT) |  | Significant correlation between SMRT and vowel identification scores (0.33) |
| Gifford et al., 2018 | CI listeners (n = 477) (Mean age at CI = 62.5 yo) (Mean duration of CI experience = 3 years) | Quick spectral modulation detection (QSMD) | CNC word recognition in quiet | Significant correlation between QSMD and word recognition in quiet (0.27) |
|  |  |  | AzBio sentence recognition in quiet | Significant correlation between QSMD and sentence in quiet (0.26) |
|  |  |  | AzBio sentence recognition with a multi-talker babble and +5 dB SNR | Significant correlation between QSMD and sentence in noise recognition (0.25) |
| DeVries & Arenberg, 2018 | CI listeners (n = 13) (Mean age at CI = 53.15 yo) (Mean duration of CI experience = 9.69 years) | Psychophysical tuning curve (PTC) | CNC word recognition in quiet | Non-significant |
| Lawler et al., 2017 | CI listeners (n = 25) (Mean age at CI = 52.64 yo) (Mean duration of CI experience = 6.6 years) | Spectral-temporally modulated ripple test (SMRT) | AzBio Sentence recognition with a +8 dB SNR | Significant correlation between SMRT and sentence recognition in noise (0.69) |
| Holden et al., 2016 | CI listeners (n = 39) (Mean age at CI = 58.9 yo) (Mean duration of CI experience = ~ 5 years) | Spectral-temporally modulated ripple test (SMRT) | CNC word recognition in quiet | Significant correlation between SMRT and word recognition in quiet (0.38) |
|  |  |  | AzBio sentence recognition in quiet | Significant correlation between SMRT and sentence recognition in quiet (0.36) |
|  |  |  | AzBio sentence recognition in 4-talker babble noise with a +8 dB SNR | Significant correlation between SMRT and sentence recognition in noise (0.36) |
|  |  |  | HINT sentence recognition in R-SPACE | Significant correlation between SMRT and sentence recognition in noise in R-SPACE (0.23) |
| Winn et al., 2016 | CI listeners (n = 19) and TH (N =10) (Mean age at CI = 54 yo) (Mean duration of CI experience = 5.81) | Consonant categorization using /ba/-/da/, /ra/-/la/ and /sa/-/ʃa/ continuum. | CNC word recognition in quiet | Significant correlation between the use of formant transition cue and word recognition (0.51) |
|  |  | Spectral ripple discrimination threshold (SRD) |  | Significant correlation between SRD and word recognition (0.44) |
| Winn & Litovsky, 2015 | CI listeners (n = 19) and TH (N = 10) (Mean age at CI = 48.31 yo) (Mean duration of CI experience = 7.31 years) | Consonant categorization using /ba/-/da/, /ra/-/la/ and /sa/-/ʃa/ continuum. | CNC word recognition in quiet | Significant correlation between word recognition scores and use of formant transition cue (0.27)  Non-significant correlation between spectral tilt and word recognition |
| Won et al., 2015 | CI listeners (n = 23) and TH (N = 10) (Mean age at CI = 47 yo) (Mean duration of CI experience = 4.3 years) | Spectrotemporal modulation detection (STM) | Korean version of Central Institute for the Deaf sentence recognition in quiet (K-CID) | Significant correlation between STM for lower spectral densities of 0.5 and 1.0 c/o with K-CID sentence recognition in quiet (0.42) even after controlling for the effect of temporal modulation detection. |
|  |  |  | Korean hearing in noise test (K-HINT) | Significant correlation between STM for lower spectral densities of 0.5 and 1.0 c/o and K-HINT (0.28) sentence recognition even after controlling for the effect of temporal modulation detection. |
| Jeon et al., 2015 | CI listeners (n = 28) (Mean age at CI = 55.68 yo) (Mean duration of CI experience = 5.25 years) | Spectral ripple discrimination threshold (SRD) | Consonant recognition in /aCa/ contexts in quiet | Significant correlation of SRD with consonant recognition in quiet (0.58) |
|  |  |  | Spondee word recognition in two-talker speech noise | Significant correlation of SRD with word recognition in noise (0.39) |
| Drennan et al., 2014 | CI listeners (n = 28) (Mean age at CI = 55 yo) (Mean duration of CI experience = 6.07 years) | Clinical spectral ripple discrimination threshold (SRD) | CNC word recognition in quiet | Significant correlation between word recognition and clinical SRD in quiet (0.58). |
| Gifford et al., 2014 | CI listeners (n = 76) (Mean age at CI = 54 yo) (Mean duration of CI experience = 4.4 years) | Quick spectral modulation detection threshold (QSMD) | CNC word recognition in quiet. | Significant correlation between QSMD and word recognition in quiet (0.66) |
| Anderson et al., 2012 | CI listeners (n = 15) (Mean age at CI = 50.46 yo) (Mean duration of experience with CI = 8.58 years) | Spectral ripple detection threshold, referred as spectral modulation transfer function (SMTF) | Sentence recognition with IEEE sentences in quiet | Significant correlation between SMTF with word in sentence recognition in quiet (0.68) |
|  |  |  | Sentence recognition with IEEE sentences in noise with 0, 5, 10, 15, and 20 dB SNR. | Non-significant correlation between SMTF and word in sentence recognition in noise |
|  |  |  | Vowel identification in /hVd/ contexts in quiet | Significant correlation between SMTF with low ripple densities and vowel recognition in quiet (0.63) |
|  |  |  | Vowel identification in /hVd/ contexts in noise with 0, 5, 10, 15, and 20 dB SNR | Significant correlation between SMTF with low ripple densities and vowel recognition in noise (0.3) |
| Anderson et al., 2011 | CI listeners (n = 15) (Mean age at CI = 50.46 yo) (Mean duration of experience with CI = 8.58 years) | Spectral ripple discrimination threshold (SRD) | Sentence recognition with IEEE sentences in quiet | Significant correlation between SRD and word in sentence recognition in quiet (0.59).  Non-significant correlation with STC bandwidth |
|  |  |  | Sentence recognition with IEEE sentences in noise with 0, 5, 10, 15, and 20 dB SNR. | Non-significant correlation with word in sentence recognition in noise |
|  |  | Spatial tuning curves bandwiths (STC bandwidths) | Vowel identification in /hVd/ contexts in quiet | Non-significant correlation between SRD and vowel recognition in quiet |
|  |  |  | Vowel identification in /hVd/ contexts in noise with 0, 5, 10, 15, and 20 dB SNR | Non-significant correlation between SRD and vowel identification in noise |
| Nelson et al., 2011 | CI listeners (n = 15) (Mean age at CI = 50.66 yo) (Mean duration of experience with CI = 5.46 years) | Forward masking spatial tuning curve (fmSTC) | IEEE Sentence recognition in quiet and in noise (speech shaped with 10 SNR) | Non-significant |
|  |  |  | Vowel (/hVd/) and consonant (/aCa/) identification in quiet. |  |
| Saoji et al., 2009* | CI listeners (n = 25) (Mean age at CI = 49.66 yo) (Mean duration of experience with CI = 2.13 years) and TH (n = 10) | Spectral modulation threshold (SMT) | Vowel and with /bVt/ context in quiet | Significant correlation between SMT and vowel identification (0.59) |
|  |  |  | consonant identification with /aCa/ context in quiet | Significant correlation between SMT and consonant identification (0.47) |
| Hughes & Stille, 2008 | CI listeners (n = 18) (Mean age at CI = 51.6 yo) (Mean duration of experience with CI = 1.83 years) | Psychophysical forward masking (PFM) | CNC word recognition in quiet | Non-significant |
|  |  |  | Phoneme recognition within CNC words in quiet |  |
|  |  |  | Sentence recognition with BKB-SIN with SNR changing from 21 dB. |  |
| Won et al., 2007 | CI listeners (n = 31) (Mean age at CI = 54.7 yo) (Mean duration of experience with CI = 3.98 years) | Spectral ripple discrimination threshold (SRD) | CNC word recognition in quiet | Significant correlation between SRD and CNC word recognition in quiet (0.25) |
|  |  |  | Spondee word recognition in in two-talker babble and steady-state noise | Significant correlation between SRD and Spondee words in steady-state noise (0.38) and two-talker babble (0.3) |
| Henry et al., 2005 | CI listeners (n = 23) and TH (N = 12) (Mean age at CI = 62.17 yo) (Mean duration of experience with CI = 3.45 years) | Spectral peak resolution threshold | Vowel recognition with /hVd/ contexts in quiet | Significant correlation between Spectral peak threshold and vowel recognition in quiet (0.27) |
|  |  |  | consonant recognition with /aCa/ contexts in quiet | Significant correlation between Spectral peak threshold and consonant recognition in quiet (0.36) |
| Henry & Turner, 2003 | CI listeners (n = 21) and TH (N = 8) (Mean age at CI = 54.9 yo) (Mean duration of experience with CI = 2.88 years) | Spectral ripple discrimination threshold (SRD) | Vowel recognition in /hVd/ contexts in quiet. | Significant correlation between the SRD and vowel identification in quiet (0.42) |
| Donaldson & Nelson, 2000 | Predominantly MPEAK CI listeners (n = 14) (Mean age at CI = 50.4 yo) (Mean duration of experience with CI = 4.9 years) and predominantly SPEAK CI listeners (n = 11) (Mean age at CI = 47.5 yo) (Mean duration of experience with CI = 4.72 years) | Electrode pitch ranking | Consonant recognition with /aCa/ context in quiet. | Significant improvement in consonant recognition with average spatial distance between electrodes above 4.5 mm (No R^2^ reported) |
| Zwolan et al., 1997 | CI listeners (n = 11) (Mean age at CI = 55.36 yo) (Mean duration of experience with CI = 3.27 years) | Electrode discrimination | Vowel and consonant recognition in /hVd/ and /aCa/ contexts | Non-significant |
|  |  |  | Monosyllabic word recognition with NU6 list |  |
|  |  |  | Phoneme in word recognition with NU6 list |  |
|  |  |  | Sentence recognition with CID Everyday Sentences |  |
| Nelson et al., 1995 | CI listeners (n = 14) (Mean age at CI = 47.92 yo) (Mean duration of experience with CI = 9.78 years) | Electrode pitch ranking | Consonant recognition with /aCa/ contexts in quiet | Significant correlation between electrode pitch ranking and consonant recognition based on place of articulation (0.38) at certain spatial separation of electrodes. |

*Saoji et al. (2009) and Litvak et al. (2007) were based on the data from the same participants. Since the data were statistically analyzed in Saoji et al. (2009) to examine the relationship between SMT and speech recognition, only their findings are included in the table.
